# Transmission thresholds for the spread of infections in healthcare facilities

**DOI:** 10.1101/2025.02.21.25322698

**Authors:** Damon J.A. Toth, Karim Khader, Christopher Mitchell, Matthew H. Samore

## Abstract

Some infections may be sustained in the human population by persistent transmission among patients clustered in healthcare facilities, including patients colonized with multi-drug-resistant organisms posing a major health threat. A nuanced understanding of facility characteristics that contribute to crossing a threshold for self-sustaining outbreak potential may be crucial to designing efficient interventions for lowering regional disease burden and preventing infections among high-risk patients. Using a mathematical model, we define the facility basic reproduction number *R*_0_, where a single facility can sustain an outbreak without ongoing importation under the threshold condition *R*_0_ > 1. We define *R*_0_ for a general model with heterogeneous patient states of susceptibility and transmissibility and with generic length-of-stay assumptions, and we provide a software package for numerical calculation of user-defined examples. We estimate *R*_0_ using published data for carbapenemase–producing Enterobacteriaceae (CPE) in longterm acute-care hospitals (LTACHs) and the effects of interventions on *R*_0_, including surveillance, pathogen reduction treatments, and length-of-stay reduction. In a simple model, *R*_0_is directly proportional to the sum of the mean and variance-to-mean ratio of the length-of-stay distribution. In intervention models, *R*_0_ depends on the moment-generating function of the length-of-stay distribution. From the CPE data, we estimated *R*_0_ = 1.24 (95% CI: 1.04, 1.45) prior to intervention. Weekly surveillance with 50% transmission reduction of detected patients alone could have reduced *R*_0_ to 0.85 (0.72, 0.98), with additional reduction if detected patients could be decolonized. Reducing the mean length of stay does not necessarily reduce *R*_0_ if the variance-to-mean ratio is not also reduced. We conclude that *R*_0_ >1 conditions plausibly exist in LTACHs, where CPE outbreaks could be sustained by patients who acquire colonization and subsequently transmit to other patients during the same hospital stay. Our findings illuminate epidemiological mechanisms producing those conditions and their relationship to interventions that could efficiently improve population health.

## 1. Introduction

Infections caused by multi-drug resistant organisms (MDROs) have limited available treatments and pose a significant health threat (1). Some MDROs have shown the ability to disseminate rapidly among patients within and between certain healthcare facilities (2), suggesting that efforts to reduce patient-to-patient transmission via interfacility coordination (3) and/or interventions targeted at high-risk settings (4, 5) could provide substantial population benefit. Results from simulations of regional outbreaks and interventions suggest that, in some scenarios, efforts to prevent transmission such as active surveillance for MDRO carriage among high-risk patients could drastically reduce regional vulnerability to explosive outbreaks that would lead to endemic presence of the MDRO (4). These results point to the possibility that rates of MDRO transmission between healthcare patients could be close to the threshold required to sustain long-lasting chains of person-to-person transmission that are characteristic of successful invasion of a novel transmissible organism in a population.

The quantitative theory of transmission thresholds has generated a rich body of research. Levels of transmission for an organism in a population relative to a threshold are often characterized by the basic reproduction number, *R*_0_, the expected total number of transmissions from a typical infected individual in an unexposed population (6). If *R*_0_ is larger than the threshold of 1, the organism can invade and produce a sustained outbreak over many generations of transmission. When above the threshold, the value of *R*_0_ can determine the theoretical level of transmission-reducing intervention effort, such as vaccination, required to control outbreaks (7).

Some studies have applied transmission threshold and reproduction number theory specifically to organisms transmitted primarily in healthcare settings, usually defining *R*_0_ as the expected number of transmissions from an admitted carrier of an organism during a single stay in a healthcare facility under specific assumptions for a particular pathogen (8-11). Others have defined this quantity as the admission reproduction number, *R*_*A*_ (12-17), or ward reproduction ratio, *R*_*w*_ (18), noting that the quantity is less than *R*_0_ when a single facility or ward stay may represent only a portion of the patient*’*s potential to transmit during the infectious period.

While the reproduction number in a healthcare facility is partly determined by properties of the organism and its typical course of colonization and infection in humans, it can also be affected by actions the facility takes to prevent patient-to-patient transmission. Of particular interest are scenarios where a facility*’*s *R*_0_ >1 for an MDRO under the facility*’*s standard transmission control efforts, but additional actions, such as active surveillance and contact precautions for detected carriers of an MDRO (19) or decolonization via pathogen reduction approaches (5, 20), could potentially move *R*_0_ below threshold.

Our aims for this work were first to define a facility reproduction number for a broad class of mathematical models and demonstrate its utility as a threshold for facility outbreak potential, and second to demonstrate our model*’*s utility in assessing intervention components that might push a vulnerable facility below threshold. For this second aim, we use data from a bundled intervention that successfully reduced infections with carbapenemase-producing Enterbacteriaceae (CPE) in long-term acute care hospitals (LTACHs) (21). Due to their composition of acutely ill patients with long length of stay, LTACHs are high-risk settings for infections with CPE and other high-priority organisms (22). Efforts to interrupt transmission could provide substantial benefit to LTACHs and other facilities linked by patient exchange (4). Therefore, understanding the role of individual intervention components for reducing transmission could be crucial to achieving maximum efficiency in regional outbreak prevention.

## 2. Methods

### 2.1 General model description

First, we defined a general class of mathematical models describing states of patients in a healthcare facility. We split the patients into different states enumerated by integer subscripts *i*, and let *x*_*i*_(*t*) be the probability that a patient is alive, not discharged, and in state *i* at time *t*after admission. Defining the column vector of probabilities **x**(*t*) containing each *x*_*i*_(*t*), we define the system of differential equations

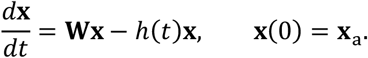

Here, **W** contains state-to-state transition rates and may also contain state-specific facility removal rates by death or live discharge. The hazard function *h*(*t*) is a facility removal rate at time *t* of the stay, where the same function applies to every state – this function can be used to calibrate a model to an overall observed patient length of stay distribution. The vector **x**_a_ is the distribution of states at admission. For this system form, we derive the formula for the equilibrium cross-sectional state distribution of facility patients, under the assumption that patients are continually admitted to the facility at a constant rate with an unchanging admission state probability distribution.

Next, for modeling the dynamics of a particular organism that can colonize or infect patients in the facility, we assume the patient states **x** are comprised of *n* susceptible states (*S*_1_,…,*S*_*n*_) and *m* colonized states (*C*_1_,…,*C*_*m*_). The colonized states represent patients carrying the modeled organism. The *m*different colonized states could represent any epidemiologically relevant differences between different groups of colonized patients, for example, states of infection at different body sites, different levels of transmissibility to other patients, different hazards of death or discharge, etc. The susceptible states represent patients not colonized by the organism. The *n* different susceptible states could be distinguished by different risks of acquiring colonization per level of exposure and/or by other relevant differences.

We use the following notation:

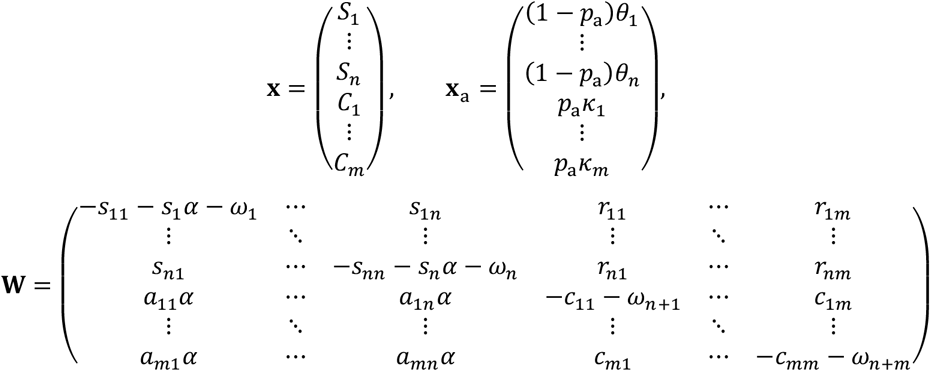

In the initial state vector **x**_a_ describing the admission state distribution, *p*_a_ is the overall probability of being in a colonized state at admission, (*θ*_1_,…,*θ*_*n*_) is the admission state distribution among the susceptible state admissions, with ∑ *θ*_*i*_ =1, and (*k*_1_, … *k*_*m*_) is the admission state distribution among the colonized state admissions, with ∑ *k*_*i*_ =1.

In the matrix **W**, the terms (*ω*_1_, …, *ω*_*n* +*m*_) in the diagonal elements are the rates of removal from the facility for patients in each state, and the other terms describe transitions from one state to another. The upper-left *n × n* portion of **W** contains elements describing the transitions out of and between the susceptible states. The *s*_*ij*_ terms are the rates of transitioning from one susceptible state to another, with *s*_*jj*_ = ∑_*i*≠*j*_ *s*_*ij*_ if *n* >1 and *s*_11_ =0 if *n* = 1. The *s*_*j*_ *α* terms are the rates of transitioning from each susceptible state to a colonized state (rates of acquisition). The *s*_*j*_ coefficients describe relative susceptibility to acquisition of patients in each susceptible state, and *α* is a baseline acquisition rate that is assumed to depend on the prevalence of patients in the colonized states, who can transmit to the susceptible patients, as follows:

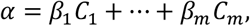

where the (*β*_1_, …, *β*_*m*_) coefficients define the relative transmissibility of patients in the different colonized states.

The lower-left *m × n* portion of **W** contains rates of transitioning into each colonized state from each susceptible state, with *s*_*j*_ = ∑ *a*_*ij*_. The lower-right *m × m* portion of **W** contains elements describing the transitions out of and between the colonized states. For the *c*_*jj*_ elements in the diagonal we have *c*_*jj*_ = ∑_*i*≠*j*_ *c*_*ij*_ + ∑ *r*_*ij*_, where the *r*_*ij*_ terms comprising the upper right *n* × *m* portion of **W** are the rates of transitioning from colonized to susceptible, i.e. the rates of clearing colonization.

For this system, we derive a formula for the facility basic reproduction number *R*_0_ for given values of the importation state distribution, state transition rates, and relative transmissibility parameters defined above, using the procedure outlined in Diekmann et al. (6). We also derive *R*_0_ formulae for a series of specific model examples.

### 2.2 Example model descriptions

#### 2.2.1 Model 1: Simple susceptible–colonized model

We specified a simple version of our general disease model with only two states and one direction of transition between states. The state probability vector **x** consists of one susceptible state (*S*_1_) that we rename *S* and one colonized state (*C*_1_) that we rename *C*. The admission state probability vector in the above notation contains *θ*_1_ = 1 and *k*_1_ = 1:

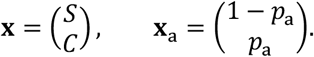

We assume that susceptible inpatients acquire colonization at rate *α*, and there are no other state transitions or removals other than removal modeled by the hazard function *h*(*t*). Thus, in the general model notation, we have *s*_1_ =1, *a*_11_ =1, *r*_11_ =0, *ω*_1_ = *ω*_2_ =0. The state transition matrix **W** is then

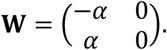

For transmission, there is only one colonized state from which transmissions can occur, so there is a single transmission parameter (*β*_1_) that we rename *β*, and the acquisition rate *α* = *βC*. The time-of-stay-dependent removal hazard *h*(*t*) is left general. The equivalent system in non-matrix form is as follows:

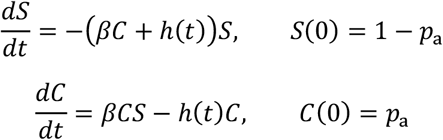

#### 2.2.2. Model 2: Clearance of colonization

To extend Model 1, we add a non-zero clearance rate *γ* of moving from colonized back to susceptible. In the general model notation, we have *s*_1_ = 1, *a*_11_ = 1, *r*_11_ = *γ, ω*_1_ = *ω*_2_ =0. The state transmission matrix **W** is then

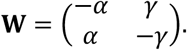

In non-matrix equation form, we have:

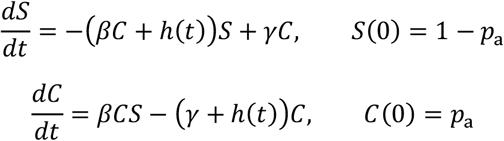

#### 2.2.3 Model 3: Clinical detection with contact precautions

For this model, we extend Model 2 to include two colonized states (*C*_1_,*C*_2_) that we rename *C* and *C*_cd_. These two colonized states represent, respectively, patients with no prior detection of their colonization and patients with a prior clinical detection. This type of model is generally used to represent the transition from asymptomatic colonization to the onset of symptomatic, invasive infection that triggers a clinical test for the causative organism. For the admission state probability vector in the above notation, we assume *θ*_1_ = 1, *κ*_1_ = 1, and *κ*_2_ = 0, i.e., all colonized patients are undetected at admission:

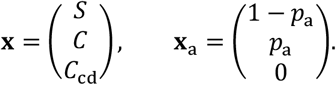

For state transitions among inpatients, we assume that susceptible patients acquiring colonization (at rate *α*) move into the undetected colonized state. Undetected colonized patients move to the susceptible state at clearance rate *γ* and move to the clinically detected state at clinical detection rate *δ*_c_, which represent a progression rate from asymptomatic colonization to confirmed clinical infection. Clinically detected patients are assumed to remain in that state until the end of the stay. Thus, in the general model notation, we have:

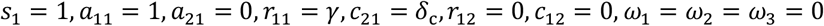

The state transmission matrix **W** is then

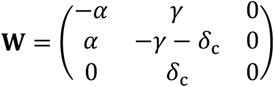

For the relative transmission rates from patients in the two colonized states, we assume that clinically detected patients transmit at a rate reduced according to the effectiveness *ε* of contact precautions: *β*_1_ = *β, β*_2_ = *β*(1 **−** *ε*). Then the acquisition rate *α* = *β*(*C* +(1 **−** *ε*)*C*_cd_).

In non-matrix equation form, Model 3 is written as follows:

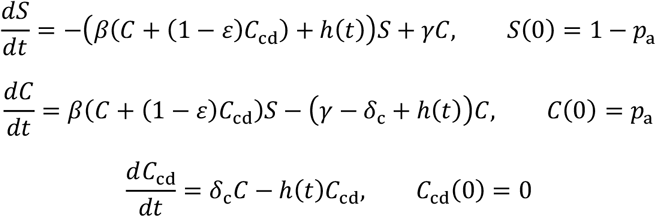

#### 2.2.4 Model 4: Active surveillance and decolonization

We extend Model 3 to include two simultaneous interventions: active surveillance to detect patients with colonization who have not been detected by clinical test, and decolonization of those detected patients via administration of a decolonizing drug or pathogen reduction agent. Patients detected by active surveillance are assumed to be placed under contact precautions in addition to receiving the pathogen reduction treatment, which increases the rate of clearance above the natural clearance rate.

The state probability vector **x** consists of two susceptible states (*S*_1_,*S*_2_) that we rename *S* and *S*_*s*d_ and three colonized states (*C*_1_,*C*_2_,*C*_3_) that we rename *C,C*_*s*d_, and *C*_cd_. The new states with subscript *“*sd*”* represent patients with a positive surveillance test earlier in the facility stay. At admission, we assume that susceptible patients never test positive, and colonized patients test positive with probability *Π*_*a*_, which incorporates the probability of receiving an admission surveillance test and the probability that the test avoids a false negative result. For the admission state probability vector in the above notation, we assume *θ*_1_ = 1, *θ*_2_ = 0, *k*_1_ = 1 **−** *Π*_*a*_, *k*_2_ = *Π*_*a*_, *k*_3_ = 0:

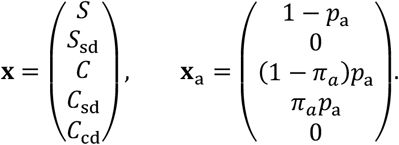

For state transitions among inpatients, we assume that undetected colonized patients become surveillance detected at rate *δ*_s_, representing mid-stay surveillance performed on patients who did not test positive at admission. Surveillance-detected, colonized patients can move to the clinically detected state at the same rate *δ*_c_ as undetected colonized patients. The new susceptible state *S*_*s*d_ can be reached by surveillance-detected, colonized patients who clear colonization during the stay. Surveillance-positive patients are assumed to remain under contact precautions for their entire remaining stay. For susceptible patients under contact precautions, we assume that those precautions reduced their rate of re-acquisition by the same factor, (1 **−** *ε*), that they reduce transmissibility. In the general model notation we have the following nonzero elements: *s*_1_ = *a*_11_ = 1, *s*_2_ = *a*_22_ = 1 **−** *ε, r*_11_ = *γ, c*_21_ = *δ*_*s*_, *c*_31_ = *δ*_c_, *r*_32_ = *γ*_d_, *c*_32_ = *δ*_c_, *ω*_1_ = *ω*_2_ = *ω*_3_ = *ω*_4_ = *ω*_5_ = 0. The state transmission matrix **W**is then

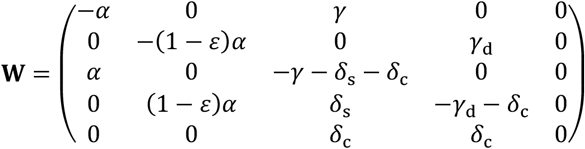

For relative transmission rates, we assume that surveillance detected, colonized patients transmit at a rate commensurate with clinically detected patients: *β*_1_ = *β, β*_2_ = *β*_3_ = *β*(1 **−** *ε*). Then the acquisition rate *α* = *β*(*C* + (1 **−** *ε*)(*C*_*s*d_ + *C*_cd_)).

In non-matrix equation form, Model 4 is written as follows:

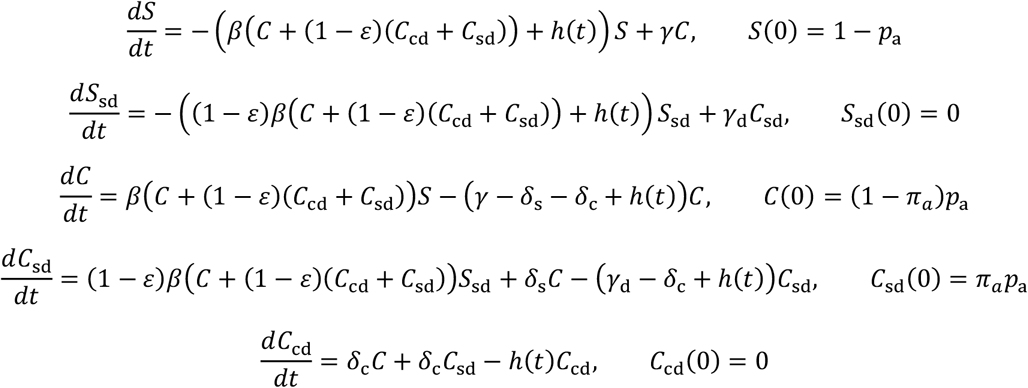

### 2.3. Calibration of Model 3 to pre-intervention CPE in LTACH

As in our prior work (5, 23), we calibrated model parameter values to data for CPE in Chicago-area LTACHs (21). Prior to intervention in those LTACHs, cross-sectional prevalence surveys revealed nearly 50% prevalence of CPE carriers, and there were significant rates of CPE clinical detection (Table 1). The facilities did not routinely perform CPE surveillance before the intervention, thus we used our Model 3 to represent the pre-intervention scenario with no active surveillance. For the facility removal (discharge or death) hazard, we used a function giving rise to a length-of-stay distribution governed by a mixture of an exponential and a gamma distribution, which in prior work we found to provide a parsimonious fit to detailed LTACH length of stay data (4). The distribution has a total of four parameters: the rate parameter of an exponential distribution, *r*_*x*_, the rate and shape parameters of a gamma distribution, *r*_*g*_ and *k*, and the portion of patients following the exponential distribution, *p*_*x*_. We calculated the unique values of these four parameters that produce a length-of-stay distribution matching the following statistics reported in the LTACH study (21): the mean, median, and interquartile range of hospital stay (Supplementary Material).

**Table 1.** Data from pre-intervention LTACHs (21)

| Value (symbol) | Data | 95% range |
| --- | --- | --- |
| Admission CPE positivity fraction ($a$) | 0.206 | (0.189, 0.223) |
| Cross-sectional facility CPE positivity fraction ($f$) | 0.458 | (0.421, 0.495) |
| CPE clinical detection rate per 1000 patient days ($d$) | 3.7 | (3.4, 4.0) |
| Days of stay: mean; 25<sup>th</sup>, 50<sup>th</sup>, 75<sup>th</sup> percentiles ($\mu$, $l_{25}$, $l_{50}$, $l_{75}$) | 33.8; 16, 28, 43 | - |

For the CPE parameters in Model 3, we assumed the CPE clearance rate *γ*=1 / (387 days) (24), contact precaution effectiveness *ε*=0.5 (25), and CPE test sensitivity = 0.85 (26, 27). Because the LTACH importation rate and crosssectional prevalence of CPE were observed and stable over time, we assumed the Model 3 system was at equilibrium, which means that the acquisition rate, *α*, was constant, making Model 3 a linear system. We solved for the value of *α* and of the progression rate to clinical detection, *δ*_c_, that produced a match to the equilibrium cross-sectional CPE carriage prevalence (scaled by the assumed test sensitivity) and CPE clinical detection incidence reported in the source LTACH study (Table 1). Then we calculated the baseline transmission rate, *β*, that produced the correct acquisition rate under the calibrated model (Table 2 and supplementary material). Finally, we used the assumed values of *γ* and *ε* and the data-calibrated values of *β* and *δ*_c_ to calculate *R*_0_ using the formula we derived for Model 3 (see Results).

**Table 2.**
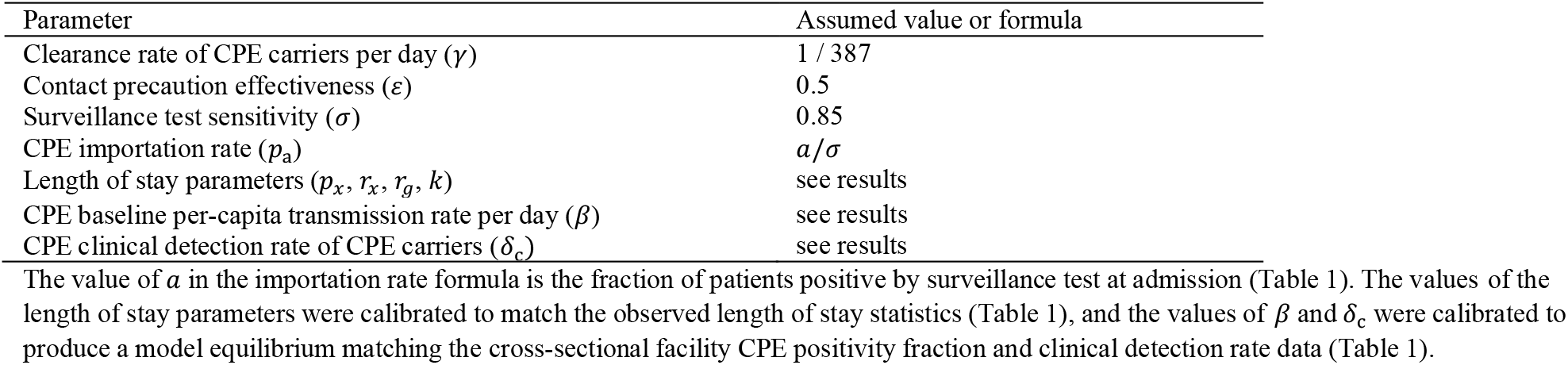
Model 3 parameters and assumptions.

We accounted for uncertainty arising from certain pre-intervention data that were used for calibration (admission CPE positivity and surveillance adherence fractions), or calibration targets (cross-sectional CPE positivity fraction and clinical detection incidence) in the model. Here, we relied on the reported uncertainty ranges from the source paper (21) (Table 1). Each value with reported uncertainty was derived from a large number of observations (21), justifying the use of a normal distribution approximation to the underlying distribution. We incorporated these assumed parameter distributions into a Latin hypercube sampling scheme (28), which produces random sets of values representative of the multi-variate distribution. We used 1000 combinations of the paper-derived values from the Latin hypercube sampling scheme to create 95% intervals for our results, including *R*_0_.

### 2.4 Application of Model 4 to intervention analysis for CPE in LTACH

We used Model 4 to investigate the potential effects of active surveillance and decolonization for CPE carriage on reducing the facility reproduction number from the pre-intervention estimates. First, we assessed the biweekly surveillance component of the intervention done in the source study (21), which in our prior work we found could largely explain the post-intervention CPE reduction they observed when 50% contact precaution effectiveness was assumed (23). The surveillance detection rate, *δ*_s_, was assumed to be the product of the targeted testing rate (once per 14 days), the reported compliance rate (0.954), and the assumed test sensitivity (0.85): *δ*_s_ ≈ 0.058 per day. We tested the effect of increasing the frequency of surveillance from biweekly to weekly and calculated the minimum frequency of surveillance testing required to make *R*_0_ < 1.

Next, we modeled the effect of combining surveillance with pathogen reduction treatment of surveillance-detected patients, represented by increasing the non-treated rate of clearing colonization among surveillance-detected patients by a factor *f* > 1, such that *γ*_d_ = *fγ*. For values of *f* = 2, 5, 10, 50, 100, and ∞ (immediate decolonization at detection), we calculated *R*_0_ for weekly and biweekly surveillance, and the surveillance rate required to achieve *R*_0_ < 1.

### 2.5 Length of stay intervention analysis

We tested the effects of interventions that would alter one or more of the parameters governing the length of stay distribution on *R*_0_, using Model 3 calibrated to LTACH CPE data (Table 1 and Table 2) as the starting point. As the overall mean length of stay decreases as either of the rate parameters *r*_*x*_ or *r*_*g*_ increases, we tested the *R*_0_-altering effects of increasing either *r*_*x*_, *r*_*g*_, or both while holding the other parameters constant, to model the effects of efforts to speed up the discharge of LTACH patients. We also tested the effects of altering the *p*_*x*_ parameter governing the portion of patients following exponential- or gamma-distributed length of stay in favor of the group with lower mean length of stay.

Finally, we chose random combinations of all 4 parameters governing the length-of-stay distribution and produced scatterplots of the resulting *R*_0_ (again using Model 3 calibrated to LTACH CPE data) vs. different length-of-stay distribution statistics. We plotted *R*_0_ against the length of stay mean, standard deviation, variance-to-mean ratio (VMR), and the sum of the mean and VMR, and we calculated Pearson*’*s correlation coefficient for the correlation between each of those statistics and *R*_0_.

## 3. Results

### 3.1 General facility basic reproduction number (R_0_) formula

For the general model described in the Methods section, we derived an expression for the basic reproduction number among facility patients. We express the result using the following components:

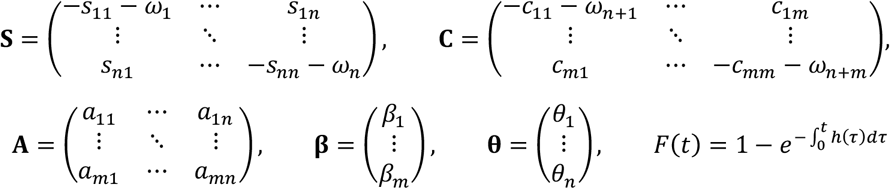

The elements in the matrices **S, A**, and **C** were defined within the matrix **W** in the Methods section. The matrix **S**describes transition rates between the susceptible states in the absence of colonized patients, and state-specific removal rates from the facility along the diagonal (*ω*_*i*_ terms). The matrix **C** contains transition rates from and between the colonized states, including state specific removal rates from the facility along the diagonal (*ω*_*i*_ terms). The matrix **A** contains the relative transition rates *a*_*ij*_ from susceptible state *j* to colonized state *i* upon acquisition. The vector ***β*** contains the transmission rates from each colonized state. The full transition rate from susceptible state susceptible state *j* to colonized state *i* is *a*_*ij*_ (*β*_1_*C*_1_ + **…** + *β*_*m*_*C*_*m*_). The vector *θ* contains the fractions of admitted, susceptible patients who are in each of the susceptible states at admission. The function *F*(*t*) is the cumulative distribution function of the facility length of stay distribution of patients who are removed via the hazard function *h*(*t*).

With these elements in hand, we derived the following result for the facility basic reproduction number *R*_0_ (Supporting Information):

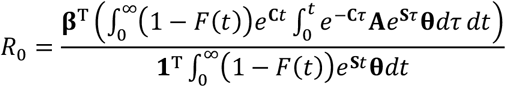

This formula is a dot product of the vector ***β***, containing the transmissibility per time of each colonized compartment, with a vector of the same length that contains the expected amount of time spent in each colonized compartment between the time of acquiring colonization in the facility and discharge or death.

We provide example models below in which *R*_0_ can be expressed symbolically, which generally occurs when the eigenvalues of matrices **S** and **C** can be expressed as formulas in terms of the model parameters. Otherwise, *R*_0_ can be calculated numerically as follows. In the numerator of the above expression for *R*_0_, the vector 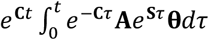 is equivalent to final *m* elements of the vector 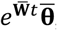, in which

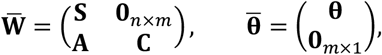

and **0**_*i×j*_ contains *i* rows and *j* columns of zeros. We numerically calculate the eigenvalues *λ*_*i*_ of 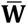 and associated eigenvectors to determine the linear combinations of 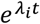 functions that comprise the elements of 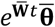.Then, integrating the product of this vector and (1 **−** *F*(*t*)) produces a vector with linear combinations of *K*(*λ*_*i*_), where *K*(*x*) = (1 **−** *M*(*x*)) ⁄ *x* and *M*(*x*) is the moment generating function of the distribution for which *F* is the cumulative distribution function.

For a model with the hazard function *h*(*t*)=0, the patients*’* rate of removal from the facility (by discharge or death) is entirely state-dependent and quantified by the *ωi* values within the matrices. Then the function *F*(*t*)=0and the basic reproduction number formula (supplementary material) reduces to:

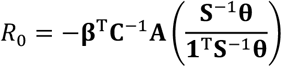

This result is valid if all patients are guaranteed to eventually be removed from the facility, i.e. there is always a nonzero probability that an inpatient will eventually reach a state *i* from which the removal rate *ω*_*i*_ >0.

We created an R package, facilityepimath [https://epiforesite.github.io/facilityepimath/] containing a function that numerically calculates *R*_0_according to the above formula, for a given set of numerical inputs for **S, C, A, β**, *θ*, and *M* (if *h*(*t*) ≠ 0).

### 3.2 Example model R_0_ formulas

#### 3.2.1 Model 1: Simple susceptible–colonized model

For this model we have:

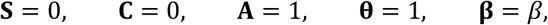

and the hazard function *h*(*t*) is left general. Substituting these values in the general formula gives

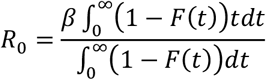

In the supplementary material we show that 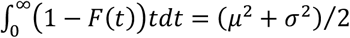, and 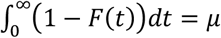, where *μ* and *σ*^2^ are the mean and variance, respectively, of the distribution for which *F*(*t*) is the cumulative distribution function. Therefore, our result is:

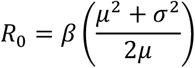

#### 3.2.2. Model 2: Clearance of colonization

For this model we have:

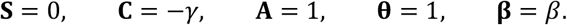

After substitution and some integration (see more detailed steps in the supplementary material), the result is:

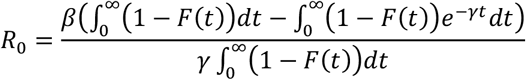

In the supplementary material we demonstrate that, for a real number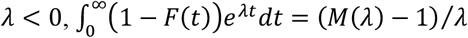, where *M* is the moment-generating function of the distribution for which *F* is the cumulative distribution function. For ease of notation, we define the function *K*(*λ*) = (*M*(*λ*) **−** 1) ⁄ *λ*. Our result for this model can then be expressed

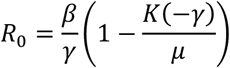

Some commonly used parametric distributions for modeling length of stay provide convenient expressions for the moment-generating function *M* within the formula for *K*. For example, for length of stay exponentially distributed with rate *r* (constant removal hazard), we have *μ* = 1 ⁄_*r*_ and *K*(*λ*) = 1 ⁄(*r* **−** *λ*), which leads to:

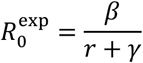

With time to removal gamma distributed with rate parameter *r* and shape parameter *k*, we have *μ* = *k* ⁄ *r* and *K*(*λ*) = ((1 **−** *λ* / *r*)^**−***k*^ **−** 1) ⁄ *λ*, which leads to

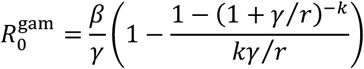

#### 3.2.3 Model 3: Clinical detection with contact precautions

For this model we have:

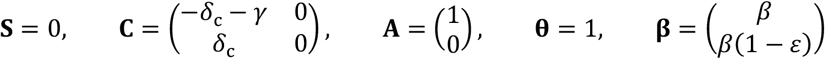

The formula for *R*_0_ contains e^**C***t*^, which, because **C** is a matrix, is the matrix exponential:

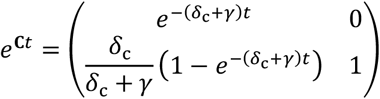

Substituting this and the other expressions in the *R*_0_ formula, we arrive at the following results after integrating and performing matrix multiplications:

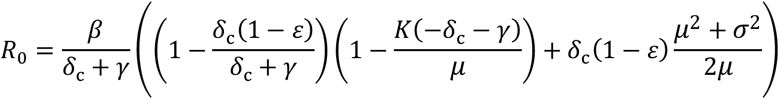

#### 3.2.4 Model 4: Active surveillance and decolonization

For this model we have:

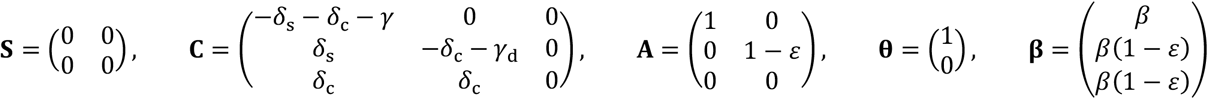

The matrix exponentials e^**S***t*^ and e^**C***t*^ can be expressed:

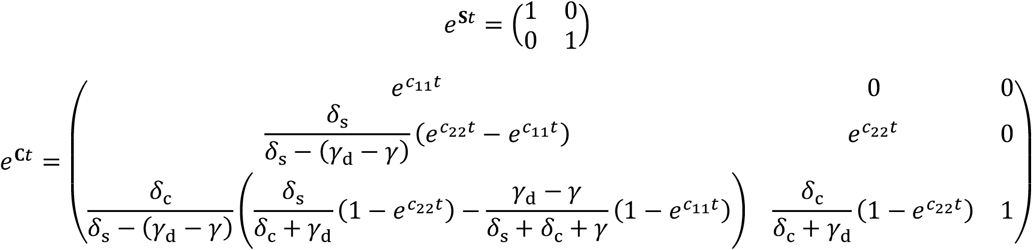

The values *c*_11_ and *c*_22_ are the first and second diagonal elements of the matrix **C**. Substituting these and the other expressions in the *R*_0_ formula, we arrive at the following result after integrating and performing matrix multiplications:

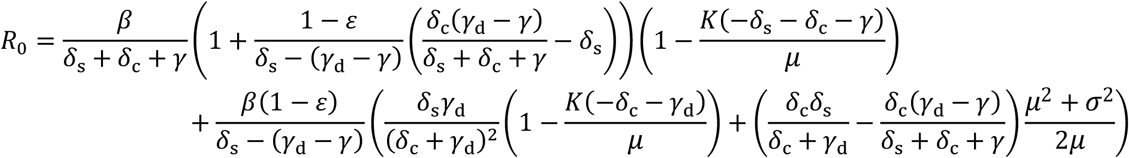

In the case that *δ*_s_ **−** (*γ*_d_ **−** *γ*) = 0, the above expression is indeterminate because of division by zero; this corresponds to the matrix **C** having a repeated eigenvalue (*c*_11_ = *c*_22_ = *δ*_s_ **−** *δ*_c_ **−** *γ*) that is degenerate, and the matrix exponential e^**C***t*^ includes eigenfunctions 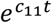 and 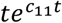. In this case, the resulting expression for *R*_0_ is as follows:

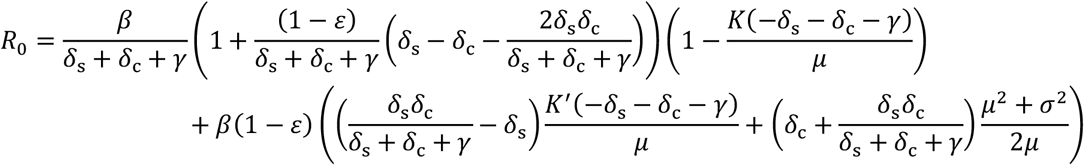

We also use simplified expressions for other special cases of the modeled intervention. First, with surveillance and contact precautions as the only intervention component, i.e. no decolonization, we have *γd* = *γ* and *R*_0_ simplifies to:

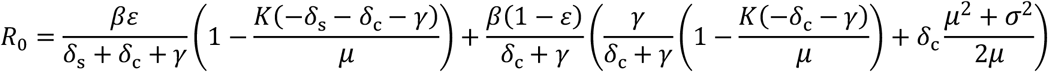

Second, when decolonization is applied to surveillance-detected patients, and the clearance effect is assumed to be instantaneous (*γ*_d_ → ∞), we can use the result from Model 2 with *γ* replaced by the sum of *γ* and *δ*_s_. That is,

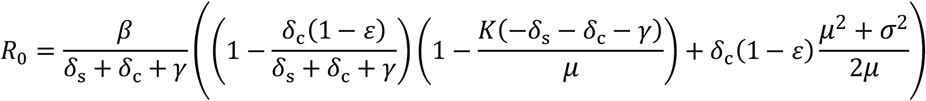

### 3.3 Calibration of Model 3 to pre-intervention CPE in LTACH

For the length of stay distribution, we assume a mixed exponential-gamma distribution, with a portion *p*_*x*_ of patients following an exponential distribution with rate *r*_*x*_ and the rest following a gamma distribution with rate *r*_*g*_ and shape parameter *k*. The following length of stay statistics reported in the LTACH study were used to estimate the 4 parameters: the mean, median, 25^th^ and 75^th^ percentile of the length of stay distribution. With 4 unknowns fit to those 4 data points (Supplementary Methods), we calculated the unique solution (Table 3).

**Table 3.** Model calibration and *R*_0_ results: CPE LTACH pre-intervention.

| Parameter | Estimated value (95% CI) |
| --- | --- |
| Exponential length of stay probability ($p_x$) | 0.580 |
| Exponential length of stay rate parameter ($r_x$) | 0.0285 per day |
| Gamma length of stay rate parameter ($r_g$) | 0.179 per day |
| Gamma length of stay shape parameter ($k$) | 5.74 |
| Base transmission rate ($\beta$) | 0.051 (0.043–0.059) per day |
| Clinical detection rate ($\delta_c$) | 0.0084 (0.0073 – 0.0097) per day |
| Facility basic reproduction number ($R_0$) | 1.24 (1.04–1.45) |

For the CPE parameters, under our assumed values for three independently based parameters (*ε*=0.5, *γ*=1 / 387 per day, *σ*=0.85), we estimated the pre-intervention values of *β, δ*_c_, and *R*_0_ from CPE data in LTACHs, using Model 3 (Table 3). Our estimate for the baseline transmission rate, *β*, was 0.051 per day (95% range 0.043 to 0.059). The estimate of rate of progression to clinical detection, *δ*_c_, was 0.0084 per day (0.0073 to 0.0097). Using those results and our formula for Model 3 above, we estimated *R*_0_ = 1.24 (1.04 to 1.45).

### 3.4 Application of Model 4 to vertical intervention analysis for CPE in LTACH

First, we estimated the effect of the biweekly CPE surveillance component of the intervention reported in the source study. Using the formula for *R*_0_ from Model 4, with *γ*_d_ = *γ* (no decolonization effect) and surveillance detection rate *δ*_*s*_=0.058 per day (once per 14 days multiplied by scaling factors for imperfect compliance and imperfect test sensitivity). With these values, and with other assumptions unchanged from the pre-intervention scenario modeled in the previous section, we estimated *R*_0_ = 0.94 (95% range 0.79 to 1.08). Increasing the frequency of surveillance from biweekly to weekly decreases the estimate to *R*_0_ = 0.85 (0.72 to 0.98). The minimum frequency of surveillance testing required to make *R*_0_ < 1 was once per 3.2 weeks (1.1 to 23 weeks) (Table 4, top row).

**Table 4:** *R*_0_ and threshold estimates for combined surveillance and decolonization intervention.

| Decolonization rate factor | Mean days to decolonization | $R_0$ with weekly surveillance (95% CI) | $R_0$ with biweekly surveillance (95% CI) | Surveillance interval needed to achieve $R_0 < 1$ (95% CI) |
| --- | --- | --- | --- | --- |
| 1 (no effect) | 387 | 0.85 (0.72, 0.98) | 0.94 (0.79, 1.08) | 3.2 weeks (1.1, 23) |
| 2 | 194 | 0.82 (0.70, 0.95) | 0.92 (0.78, 1.06) | 3.6 weeks (1.4, 25) |
| 5 | 77 | 0.76 (0.64, 0.87) | 0.87 (0.73, 1.00) | 4.5 weeks (2.0, 30) |
| 10 | 39 | 0.68 (0.58, 0.79) | 0.81 (0.69, 0.93) | 5.6 weeks (2.7, 35) |
| 50 | 8 | 0.49 (0.42, 0.56) | 0.66 (0.56, 0.76) | 8.4 weeks (4.5, 48) |
| 100 | 4 | 0.43 (0.37, 0.50) | 0.62 (0.52, 0.71) | 9.2 weeks (5.0, 51) |
| ∞ (immediate) | 0 | 0.36 (0.31, 0.41) | 0.56 (0.47, 0.64) | 10.3 weeks (5.7, 56) |

Next, we modeled the effect of combining surveillance with decolonization via pathogen reduction treatment of surveillance-detected patients, represented by increasing the non-treated rate of clearing colonization among of surveillance-detected patients by a factor *f* > 1, such that *γ*_d_ = *fγ*. For different values of *f*, we calculated *R*_0_ for weekly and biweekly surveillance, and the surveillance rate required to achieve *R*_0_ < 1 (Table 4). For example, with *f* =5 (five-fold increase in clearance rate due to the pathogen reduction treatment), the *R*_0_ estimate with biweekly surveillance was 0.87 (0.73 to 1.00), an 8% decrease from the estimate of 0.94 with no decolonization effect. The upper bound of the decolonization factor (*f* → ∞, or immediate clearance upon detection) achieves *R*_0_ = 0.56 (0.47 to 0.64) with biweekly surveillance. With immediate clearance, a surveillance interval of once per 10 weeks (6 to 56 weeks) could potentially achieve *R*_0_ < 1.

The code producing the results in Table 3 and Table 4 is available as an installed file in our facilityepimath R package [https://github.com/EpiForeSITE/facilityepimath/tree/main/inst].

### 3.5 Length of stay intervention analysis

The parameters of Model 3 governing removal (death or discharge) rates of patients were calibrated to LTACH length of stay statistics (Table 1), giving rise to an overall length of stay distribution with mean *μ*=33.8 days and standard deviation *σ* = 28.1 days. With the mixture model we used to govern removal, a fraction *p*_*x*_ =0.580 of admitted patients follow an exponentially distributed length of stay (constant discharge-or-death hazard), with mean and standard deviation *μ*_*x*_ = *σ*_*x*_ = 35.1 days. The remaining patients follow gamma-distributed length of stay with shape parameter *k* >1 (meaning the removal hazard increases with time of stay), which gives a length of stay distribution with mean *μ*_*g*_ = 32.0 days and standard deviation *σ*_*g*_ = 13.4 days. Thus, the patients following the exponential distribution have a slightly higher mean and much higher variance in length of stay than the other group. We refer to the patients following the exponentially distributed length of stay as the high-variance patients and the patients following the gamma-distributed length of stay as the low-variance patients.

We tested the effects of interventions that alter different length of stay parameter values on *R*_0_ for Model 3 calibrated to pre-intervention LTACH CPE data (Figure 1). We chose four interventions that decrease the mean length of stay from the baseline pre-intervention 33.8 days. First, when increasing the discharge rate of all patients (i.e., increasing the rate parameters *r*_*x*_ and *r*_*g*_ by the same factor), *R*_0_ decreases and crosses the *R*_0_ < 1 threshold as the mean length of stay drops below about 26 days (Figure 1A). Second, when increasing the discharge rate *r*_*x*_ of the high-variance patients only, *R*_0_ decreases below 1 as the mean length of stay drops to about 28 days (Figure 1B). Third, when increasing the discharge rate *r*_*g*_ of the low-variance patients, *R*_0_ does not decrease below 1.2 and in fact begins increasing as the mean length of stay drops below about 29 days and lower (Figure 1C). Fourth, when decreasing the fraction *p*_*x*_ of patients following the high-variance exponential distribution, *R*_0_ drops rapidly and falls below 1 as the mean length of stay drops below about 32.6 days. When *p*_*x*_ = 0 (all patients follow the low-variance gamma distribution with mean 32.0 days), *R*_0_ = 0.88.

**Figure 1.**
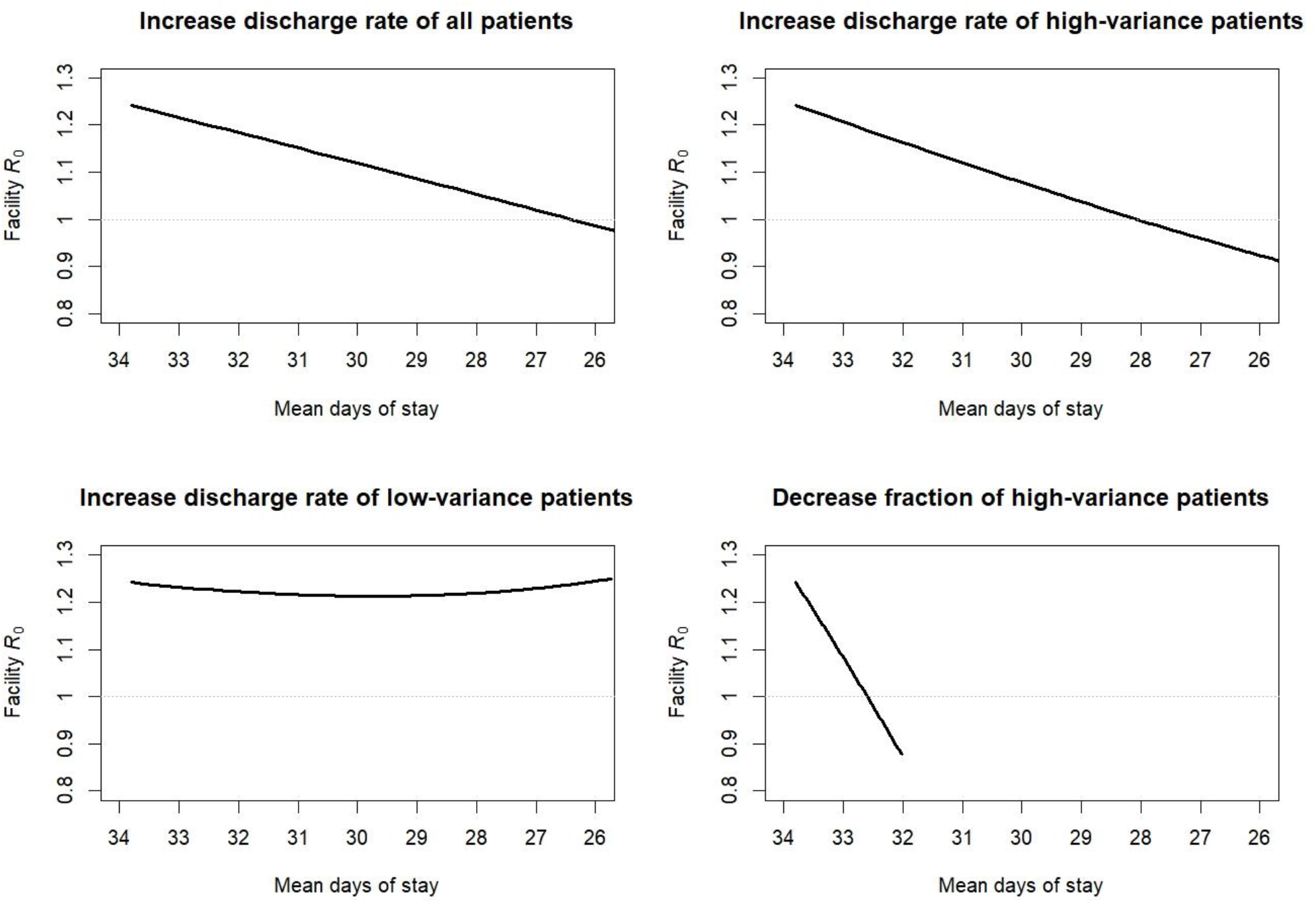
Effect of length-of-stay interventions on facility basic reproduction number Calculations of the facility reproduction number, *R*_0_, for Model 3 with the following fixed values calibrated to pre-intervention CRE data in longterm acute care hospitals (Table 3): transmission rate *β* = 0.051 per day, clinical detection rate *δ*_c_ = 0.0084 per day, colonization clearance rate *γ* = 1 / 387 per day, and contact precaution effectiveness *ε* = 0.5. Length of stay was governed by a 4-parameter mixed exponential–gammadistribution, and specific parameters were varied from the baseline values (Table 3) in a direction that decreased the overall mean length of stay from left-to-right in each of the sub-figures, while holding all other parameters constant. Upper-left: both the exponential rate parameter *r*_x_ and the gamma rate parameter *r*_g_ were simultaneously increased by the same factor from their baseline values. Upper-right: the exponential rate parameter *r*_x_ was increased from its baseline value. Lower-left: the gamma rate parameter *r*_g_ was increased from its baseline value. Lower-right: the fraction of patients following the exponential distribution *p*_x_ was decreased from its baseline value.

As the results in Figure 1 demonstrate that the mean length of stay did not necessarily correlate with *R*_0_, we tested the relationship between *R*_0_ and other statistics for the length of stay distribution (Figure 2). The dots in the scatterplot represent the results from randomly chosen combinations of the four length-of-stay parameters *p*_*x*_, *r*_*x*_, *r*_*g*_, and *k*, each picked from independent uniform distributions ranging from plus-or-minus 40% of the values from the model calibrated to the LTACH data (Table 3). For each of the random set of parameter values, we calculated *R*_0_ and the mean, standard deviation, variance-to-mean ratio (VMR) and the sum of the mean and VMR. The latter statistic was chosen because of its appearance in our result for the *R*_0_ formula for Model 1.

**Figure 2.**
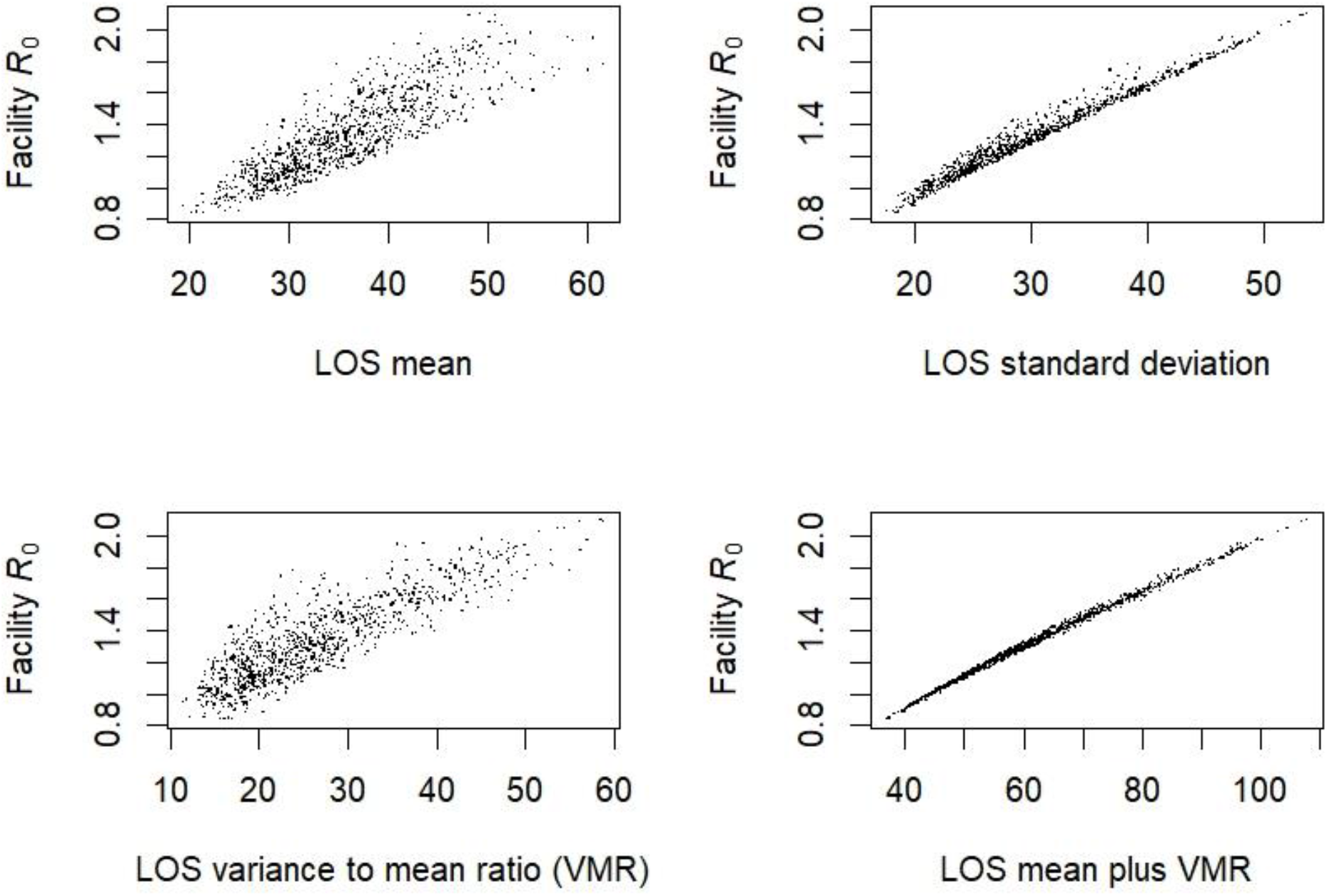
Scatterplot of facility basic reproduction number vs. length of stay (LOS) distribution statistics Calculations of the facility reproduction number, *R*_0_, for Model 3 with the following fixed values calibrated to pre-intervention CRE data in longterm acute care hospitals (Table 3): transmission rate *β* = 0.051 per day, clinical detection rate *δ*_c_ = 0.0084 per day, colonization clearance rate *γ* = 1 / 387 per day, and contact precaution effectiveness *ε* = 0.5. The dots represent *R*_0_ results for 1000 sets of values for each of the four parameters governing the mixed exponential–gamma length of stay distribution, drawn from four independent uniform distributions ranging from 60% to 140% of the calibrated values in Table 3. Each subplot displays the same 1000 *R*_0_ results, plotted against different statistics of the length of stay distribution for each set of length-of-stay parameters.

The length of stay mean produced a weaker correlation with *R*_0_ (Pearson*’*s correlation coefficient PCC = 0.857) compared to the correlation between the standard deviation and *R*_0_ (PCC = 0.986). The VMR produced PCC = 0.860, and the sum of the mean and VMR produced the strongest correlation with *R*_0_, with PCC = 0.997 and the relationship appearing nearly linear (Figure 2).

The code producing Figures 1 and 2 is available as installed files in our facilityepimath R package [https://github.com/EpiForeSITE/facilityepimath/tree/main/inst].

## 4. Discussion

This study provides a flexible, quantitative framework for calculating the basic reproduction number *R*_0_ for a wide range of mathematical models of transmissions among patients admitted to healthcare facilities. We showed that such calculations can provide actionable insights for understanding the factors leading to certain facilities reaching *R*_0_ > 1 for high-priority pathogens, creating a setting where a single patient importation can lead to a sustained outbreak. We demonstrated that one set of high-risk facilities plausibly produced *R*_0_ >11 for CPE, a multi-drug-resistant organism of high concern, and analyzed models that provided nuanced insights into alternate interventions that could reduce *R*_0_ in such facilities to below threshold.

A facility with *R*_0_ > 1 is at risk not only of explosive outbreaks among its own patients, but also for acting as a transmission amplifier within a regional healthcare network (29). Patients needing care in a high-risk (and high *R*_0_) facility like an LTACH may be likely to subsequently require care in other hospitals or long-term care facilities, and thus they pose risk of continued spread of pathogens they had acquired in the LTACH. Prior simulation studies of CPE transmission among patients in a regional network of healthcare facilities, including short-stay hospitals, nursing homes and LTACHs, demonstrated that interventions including efforts focused on detecting carriage among LTACH patients could dramatically reduce CPE infections over the whole region (4).

Those prior findings of a dramatic decrease in simulated CPE infections due to LTACH-based intervention are consistent with a theoretical result we produced here: realistic rates of surveillance for CPE carriage and contact precautions for detected CPE carriers in an LTACH can plausibly move *R*_0_ from above to below the critical threshold of 1. Crossing that threshold can explain the disproportionately high benefit achieved by a moderate increase in mitigation effort observed in simulation studies. A patient population health benefit that increases non-linearly with increasing intervention cost can also manifest in a highly efficient, cost-saving resource allocation, as predicted by a health economic analysis of a model of a CPE decolonization intervention in LTACHs (5).

Our result for the effect of CPE surveillance on reducing facility *R*_0_ was determined by the effectiveness of contact precautions to reduce transmission from detected carriers and rate of mid-stay surveillance only. The rate of *admission* surveillance does not appear in the *R*_0_ formula, because we defined *R*_0_ as the expected number of transmissions from a patient who acquires CPE after admission, between the time of acquisition and discharge or death. While admission surveillance can reduce the chances that otherwise undetected CPE importers will set off a chain of transmissions, nothing short of perfect admission detection and 100% effective contact precautions could entirely prevent a within-facility transmission from occurring. If any transmissions occur in a facility with *R*_0_ > 1 by our definition, admission surveillance would not affect continuing circulation of CPE among in-patients who were each non-carriers at admission.

Thus, our finding provides a potentially important insight for facilities considering implementing a surveillance intervention: testing patients during the middle of their stay is the critical component that reduces the chances of a facility being a transmission amplifier. A caveat to this insight is that a broader definition of *R*_0_ would cover a patient*’*s entire period of colonization, which might include multiple different stays at healthcare facilities. Admission surveillance, particularly for patients having recently stayed at the same or another high-risk facility, would help reduce *R*_0_ under that extended model.

For the single-stay facility model, we showed that the biweekly rate of surveillance for CPE reported in a Chicago area LTACH intervention (21) may have been sufficient to reduce *R*_0_ from above to below threshold, assuming 50% effectiveness of contact precautions at reducing transmission from detected patients, 95% adherence to surveillance testing, and 85% sensitivity of the test. Our *R*_0_ formula provides a quick method for testing sensitivity of *R*_0_ estimates to those assumptions as well as the rate of surveillance as an intervention decision variable.

In addition to contact precautions for identified colonized patients, the pathogen reduction intervention we tested represents the administration of treatment to detected carriers that would decolonize the patient or act to increase the rate of clearance by some mechanism, a class of treatments that has recently garnered attention (20). For organisms like CPE and other pathogenic bacteria that colonize the gut, treatments with potential pathogen reduction agents are in early stages of conception, development, testing, or approval, such as selective decontamination of the digestive tract (30), bacteriophages (31), or probiotic treatments that could fortify the human microbiome*’*s natural ability to clear harmful colonization (32). Our prior modeling study demonstrated the potential for high cost-effectiveness that a drug with rapid decolonization effects could have when administered in a high-transmission facility to even a small portion of colonized patients (5). Our results here bolster the theoretical underpinnings of that prior result and can help identify scenarios and settings where new drugs could help achieve transmission threshold effects that produces a disproportionately high health benefit. Our model combining surveillance and decolonization demonstrates a synergistic effect of the two strategies when the drug can achieve clearance rapidly.

As with any epidemiological modeling study attempting to project intervention effects in a real-world setting, our results depend on assumptions that may not be accurate. While we matched our assumptions to published data wherever possible, some key parameter value assumptions did not have strong data-based justification. For example, the relative transmissibility of detected CPE carriers compared to undetected carriers in hospitals is unknown, as the transmitter to an acquisition is rarely identified definitively during CPE outbreaks. In addition, given that the decolonization drug explored in our model is hypothetical, modeling its effects relative to the also-uncertain natural clearance rate is largely based on guesswork. Nevertheless, our equation-based results allow for quick exploration of alternate assumptions, and such sensitivity analyses in this and prior work (5, 23) suggest that our findings for these intervention effects are largely robust.

Another major finding of this study is a characteristic property of the length of stay distribution of facility patients that largely governs its relationship to *R*_0_. Namely, *R*_0_ was directly proportional to the ratio of the *second raw moment* of the length of stay distribution to the mean of the distribution. The second raw moment is the average of the squares of each individual length of stay and is also equal to the sum of the variance and the square of the mean. Thus, the characteristic ratio can also be expressed as the sum of the variance and the variance-to-mean ratio of the distribution. This is an exact result for Model 1, where colonized patients remain colonized from the time of acquisition to discharge or death with no change in their transmissibility. For more complicated models, the proportionality is not exact, as the *R*_0_ formulas depend on higher moments of the length of stay distribution via the moment-generating function. But we showed in Figure 3 that the characteristic ratio was nevertheless an excellent predictor of changes to *R*_0_ due to changes in a complicated length of stay distribution model.

Intuition as to why the square of lengths of stay is a characteristic property governing facility *R*_0_ lies in understanding the length of stay of a typical patient who might acquire colonization while they are an in-patient at a healthcare facility. If a random patient residing in the facility on a given day becomes colonized, their time remaining in the facility before discharge or death is the characteristic time duration governing their subsequent potential to transmit to others. Calculating the average length of stay of a cross-section of patients in the facility at a given time is different than calculating the average length of stay of a set of admitted patients, because longer-stay patients are overrepresented in the cross-section due to their taking up facility bed spaces for longer. Specifically, in a sample of patients in the facility at a given time, patients with a longer length of stay are more likely to be sampled, in direct proportion to their length of stay. For example, a patient with a length of stay twice as long as another was twice as likely to be in the facility on the day of sampling. Therefore, the mean length of stay of the patients sampled is a weighted mean of the actual lengths of stays of all admitted patients, where the weights are the length of stay. The weighted mean of lengths of stay weighted by length of stay reproduces the characteristic quantity: the mean of the square of length of stay divided by the mean.

This finding suggests that measures identifying each of the first two moments (e.g. mean and variance or mean and standard deviation) of the length of stay distribution are important for characterizing facility outbreak risk, and both should be more regularly calculated and considered by facilities undergoing transmission studies. Reported facility length of stay statistics often include only the mean and/or quantiles such as the median or inter-quartile range, which fail to uniquely identify the second moment.

Furthermore, we showed that the effect of efforts to reduce lengths of stay as a transmission risk-reduction intervention are not always well characterized by the intervention*’*s effect on the mean. If the mean length of stay is reduced in a way that simultaneously increases the variance-to-mean ratio, it is possible that facility outbreak risk could increase, as measured by *R*_0_. For LTACHs and similar facilities that specialize in care for high acuity patients, it might be possible to characterize the condition and care needs of incoming patients to estimate not only their expected length of stay, but also the uncertainty (i.e. variance) in days of care they will require. If a facility can control the composition of admitted patients and/or target length of stay reduction efforts on certain groups, efforts to reduce the fraction of the high-variance group of patients or to increase their discharge rates would be favored for reducing facility outbreak risk.

We have provided an R package, facilityepimath, that includes a function to calculate *R*_0_ numerically, using the general equation we derived, for a wide variety of models. The package contains example code reproducing the results for the models we investigated in this manuscript, as well as example demonstrations of how other users can use the function to calculate *R*_0_ for any model that fits our broad general framework, including models with nuanced assumptions governing facility length of stay. The package also provides other functions, e.g. to calculate the equilibrium of a model for a given, constant rate of importation of infectious individuals to the facility, which is useful for calibrating models to observed levels of admission and cross-sectional prevalence, as done in this work, and for assessing the impacts of transmission-reducing interventions under ongoing importation (5, 23).

While our formula applies to a wide range of possible models, there are limitations to what can be assumed. For contact assumptions affecting transmission, the model form requires that all susceptible compartments have the same relative exposure to individuals in different colonized compartments. Therefore, our results do not apply to segregated contact models in which, for example, the compartments represent patients in different wards of a healthcare facility with different within-ward vs. cross-ward contact rates. Our model also does not include the effects of transmission during an episode of infectiousness extending beyond a single facility stay. Extensions to multiple facility stays via readmission to the same facility and transmission in multiple different facilities or in the community outside a healthcare facility are potential subjects of future work.

In conclusion, we have derived a novel, general mathematical result for theoretical quantities that have important implications for study and control of high-priority infectious disease health threats. Our findings for healthcare facility transmission models could be applicable to other at-risk settings with similarly rapid population turnover. Locations with above-threshold transmission conditions can pose a significant risk to population health, while at the same time presenting opportunities for targeted interventions that are highly efficient. Seeking a nuanced understanding of those conditions and their relationship to intervention mechanisms should be a component of strategic planning for public health efforts.

## Data Availability

All data produced are available online at https://github.com/EpiForeSITE/facilityepimath/

## Acknowledgements

The authors acknowledge Prabasaj Paul for his role in inspiring this work and for helpful insights during its development.

## Supplementary Methods and Results

### Length of stay model calibration to LTACH data

We assume that patient length of stay is a random variable with cumulative distribution function *F*(*t*). The source LTACH study (21) provides the mean and 25^th^, 50^th^, and 75^th^ percentiles of the overall length of stay distribution (*μ, l*_25_,*l*_50_,*l*_75_).

For fitting Model 3 to data, we have four equations constraining the length of stay distribution:

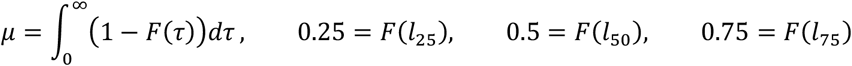

We assume a mixed exponential-gamma distribution for the length of stay distribution, with a portion *p*_*x*_ of patients following an exponential distribution with rate *r*_*x*_ and the rest following a gamma distribution with rate *r*_*g*_ and shape parameter *k*. For this distribution:

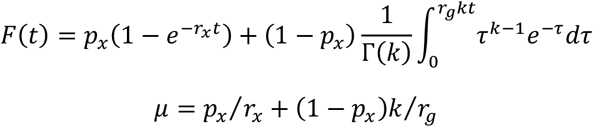

We apply these two functions to the four equations above and numerically solve for the four unknown parameters *p*_*x*_, *μ*_*x*_, *μ*_*g*_, and *k*.

From the LTACH study (21), we have *μ* = 33.8 days, *l*_25_ = 16 days, *l*_50_ = 28 days, and *l*_75_ = 43 days, which leads to:

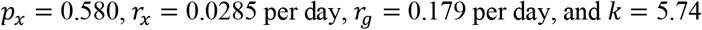

### Equilibrium calibration to LTACH CPE data

As described in the main text, we calibrated the patient acquisition rate *α* at equilibrium and the per-capita clinical detection rate *δ*_c_ of a colonized patient to data describing the cross-sectional prevalence of CPE carriage and the overall CPE clinical detection incidence rate observed at the LTACHs (21). We assumed that Model 3 (main text) was at equilibrium with the observed constant CPE admission positivity rate. Therefore, we required a formula for the equilibrium prevalence of the three patients states of Model 3 at the given initial conditions (admission states). Here we derive a formula for the equilibrium of a general model, then apply the formula to Model 3.

Let **x**(*t*) be the vector of probabilities that a patient is alive, not discharged, and in each state *x*_*i*_ at time *t*after admission. We have the system

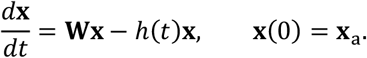

Here, **W** contains state-to-state transition rates and state-specific facility removal rates during the stay. The hazard function *h*(*t*) is the removal rate at time *t* of the stay, where the same function applies to every state. The vector **x**_a_ is the distribution of states at admission. The solution to this equation is expressed

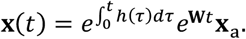

If we express the hazard function as that of a probability distribution: *h*(*t*) = *f*(*t*) ⁄ (1 **−** *F*(*t*)), where *f* and *F* are the probability density function and cumulative distribution function of the same distribution, then we can express **x**(*t*) as

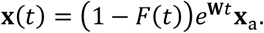

If there are no discharge or death rates within the **W** matrix, then *F* is the cumulative distribution function of the facility length of stay distribution.

If we assume that patients are continually admitted to the facility at a constant rate of qpatients per time, *ad infinitum*, then the expected number of patients in the facility between stay-time *t*_1_ and *t*_2_ in each state is 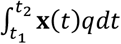. So, the expected total number of inpatients in each state is given by the vector

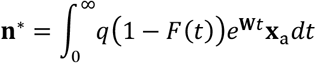

And the expected total number of patients in the facility is **N**^∗^ = **1**^T^**n**^∗^. Then, the equilibrium cross-sectional state distribution within the facility is **x**^∗^ = **n**^∗^ ⁄ **N**^∗^, or

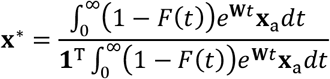

To evaluate the integral in this expression, we must consider the integral 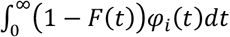 for the eigenfunctions φ that comprise the elements of the matrix exponential e^**W***t*^. If the state dependent removal rates *ω*_*i*_ are all nonzero, the eigenvalues *λ*_*i*_ of **W** are negative and real. The eigenfunctions associated with non-degenerate eigenvalues then take the form 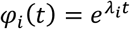. We use integration by parts:

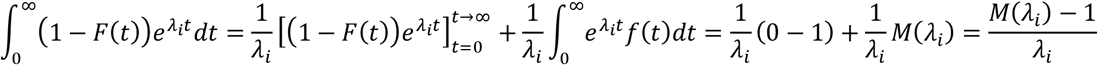

Here, *f* and *M* are the probability density function and moment-generating function of the distribution associated with the removal hazard, respectively. For ease of notation, we define the function

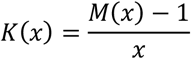

If there are any repeated, negative eigenvalues that are degenerate, there would be eigenfunctions of the form 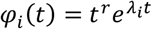 for integer *r* ≥1. We note that if 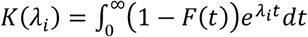 then 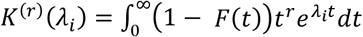, where *K*^(*r*)^ is the *r*th derivative of *K*, defined above.

Finally, for models where any state-dependent removal rates *ω*_*i*_ are set to zero, there may be an eigenvalue of **W** equal to zero, in which case there will be at least one eigenfunction *φ*_*i*_(*t*) that is constant. Then there will be a component of the 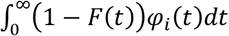 expression equal to the mean *μ* of the distribution for which *F*(*t*) is the cumulative distribution function, as 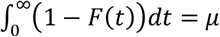.

In summary, the vector 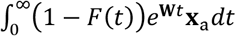 is constructed by calculating e^**W***t*^**x**_a_ and replacing the 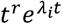 terms in each vector element with *K*^(*r*)^(*λ*_*i*_) and the constant terms with *μ*.

For Model 3, we have the following components:

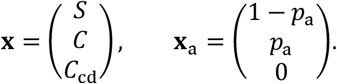

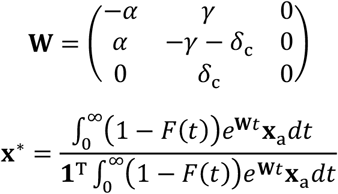

To calculate the matrix exponential e^**W***t*^ we require the eigenvalues of **W**. The first two eigenvalues are the eigenvalues of the upper-left 2-by-2 submatrix, and the third eigenvalue is 0. The first two eigenvalues *λ*_1_ and *λ*_2_ are:

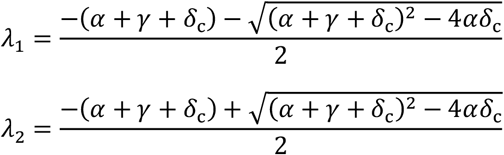

Then, the matrix exponential is:

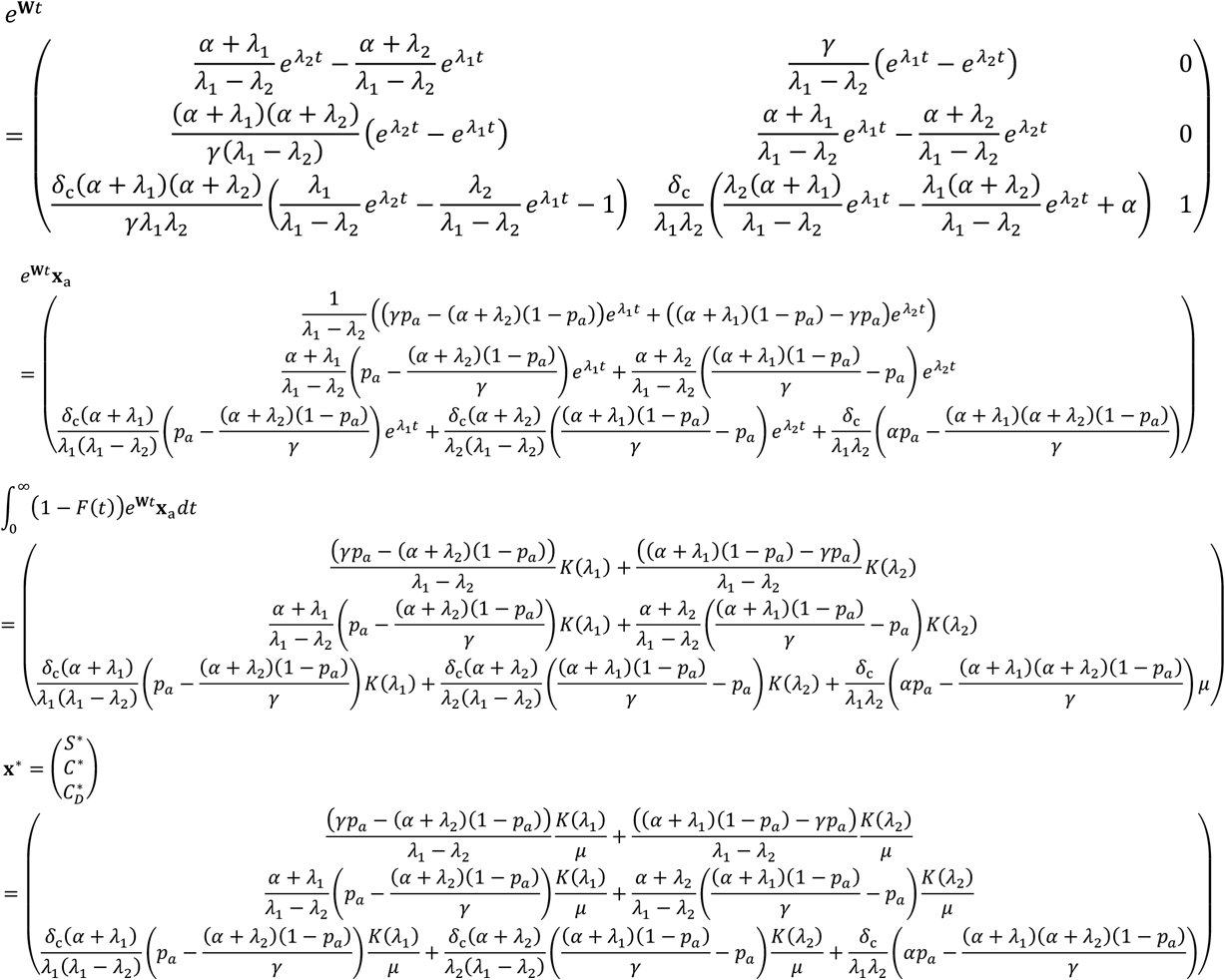

For the function *K* we require the moment generating function *M* of the mixed exponential–gamma distribution that was calibrated to the LTACH length of stay distribution as described above:

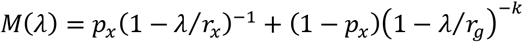

Then,

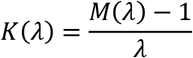

Using the above formulas, we solved for the values of *α* and *δ*_c_ that produce an equilibrium result that meets the following two criteria:

1. The cross-sectional fraction of patients who are in one of the colonized states at equilibrium is the cross-sectional positivity *f* divided by the assumed surveillance test sensitivity *σ*:

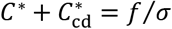
2. The clinical detection rate in the facility at equilibrium is *d*:

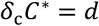

The values of *f, σ*, and *d*, as well as the other parameters aside from *α* and *δ*_c_ within the formulas for *C*^∗^ and 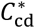 above are determined by the data and/or by assumption as described in Tables 1 and 2 in the main text. Therefore, we have two equations to solve simultaneously for the unique solution of *α* and *δ*_c_.

### Facility basic reproduction number formula derivation

For the general system defined in the main text, we will show that the facility basic reproduction number

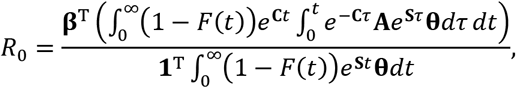

Where

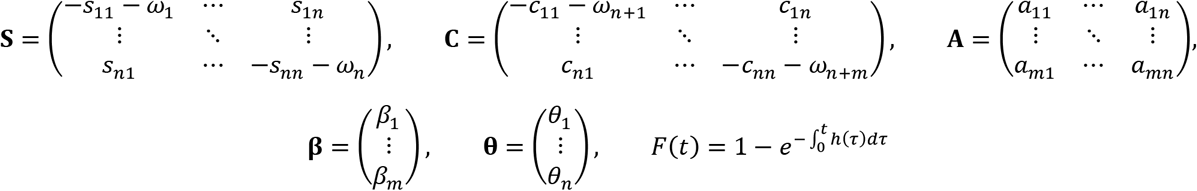

To derive our equation we show that it is consistent with a *“*basic reproduction ratio*”* formula in Diekmann et al (6). Their definition depends on a measure of an individual*’*s *“*h-state*”, η*, just prior to acquiring colonization. First, they define the quantity *S*(*η*): the distribution of h-states among susceptible individuals at equilibrium in the absence of any colonized patients. Then they define the quantity A(*τ,ξ,η*), which is the expected infectivity of an individual who was in h-state *η* just prior to acquisition time *τ* ago, toward a susceptible individual currently in h-state *ξ*.

In our model, the h-state is characterized by both the individual*’*s susceptibility compartment, *s* ∈ (1,…,*n*), and time since admission, *t* ∈ (0,∞). So we have *S*(*η*) = *S*(*s*_*η*_,*t*_*η*_) and A(*τ,ξ,η*) = A(*τ,s*_*ξ*_, *t*_*ξ*_, *s*_*η*_,*t*_*η*_). The time-since-admission component is only relevant for the infectivity of the currently colonized individual represented by *η*, whose probability of removal (death or discharge) from the facility (thus no longer infective toward other facility patients) between *t*and *t*+*τ* might depend on *t*. However, the infectivity toward the susceptible individual represented by *ξ* depends only on that individual*’*s current compartment *s*, via the susceptibility factor ψ_*s*_ (the probability of being removed while susceptible will be incorporated in the *S* function). Therefore, *A*does not depend on *t*_*ξ*_, and for simplicity of notation we let *t*= *t*_*η*_ and seek expressions for the functions *S*(*s*_*η*_,*t*) and A(*τ,s*_*ξ*_, *s*_*η*_,*t*).

The infectivity *A* will be the susceptibility ψ_*sξ*_ times the sum of infectivity values *β*_*c*_ of each possible colonized compartment *c*, weighted by the probabilities *p*(*c,s*_*η*_, *t,τ*) of being in colonized compartment *c* time *τ* after acquisition at stay-time *t*from compartment *s*_*η*_ and still in the facility. So we have:

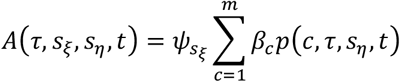

This expression for *A* satisfies a separability condition in Diekmann et al., i.e. it can be written as a product of two functions

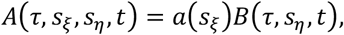

Where

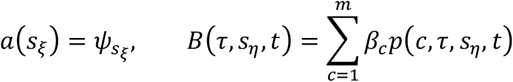

Therefore, the following formula from Diekmann et al. for the basic reproduction ratio applies:

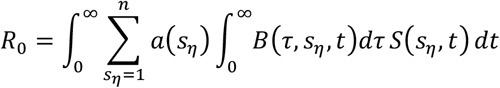

For simplicity we let *s*=*s*_*η*_ and specify the *R*_0_ formula as

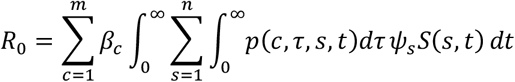

First, from our equilibrium results above we have that *S*(*s,t*) is the *s*th element of the column vector:

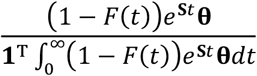

Next, we show that *p*(*c,τ,s,t*) is the element in the *c*th row, *s*th column of the matrix

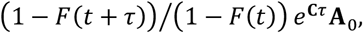

where **A**_0_ is the matrix **A** above with each column scaled by its sum. The *s*th column of **A**_0_represents the distribution of colonized compartments initially entered when acquiring from compartment *s*, and e^**C***τ*^ projects the initial compartment forward *τ* time units according to the colonized state transition dynamics embedded in **C**. The expression (1 **−** *F*(*t* + *τ*)) ⁄ (1 **−** *F*(*t*)) is the probability that removal does not occur before time-of-stay *t* + *τ* given that the stay was longer than *t* (the acquisition time).

So 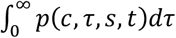 is the element in the *c*th row, *s*th column of the matrix

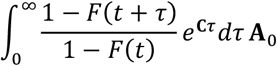

Noting that the sum of column *s* of the matrix **A** is ψ_s_, we have that 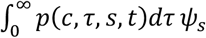 is the element in the *c*th row, *s*th column of the matrix

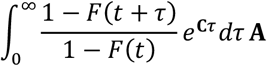

Substituting into the *R*_0_ formula above, we have

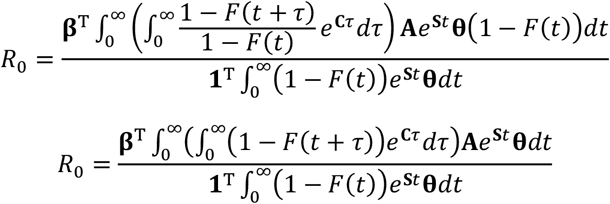

We use the substitution *z* = *τ* + *t* and rearrange terms to get

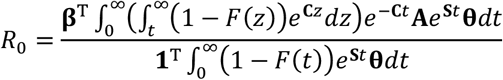

Integrate the numerator by parts using the following substitutions:

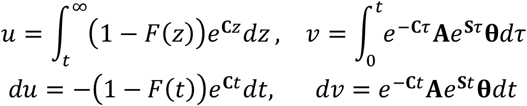

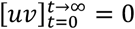,so

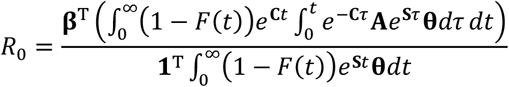

In the numerator of this expression for *R*_0_, the integral with respect to *t* can be calculated similarly to integral in the denominator, discussed above. I.e., if all state-dependent removal rates are nonzero, the elements of the vector 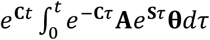 are linear combinations of 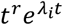 functions with negative eigenvalues can be calculated similarly to integral *i* of the **S**and **C** matrices and integer *r* ≥0. Then, integrating the product of this vector and (1 **−** *F*(*t*)) produces a vector with linear combinations of *K*^(*r*)^(*λ*_*i*_), where *K*(*x*) = (1 **−** *M*(*x*)) ⁄ *x* and *M*(*x*) is the moment generating function of the distribution for which *F* is the cumulative distribution function. If there are state-dependent removal rates equal to zero, then the vector 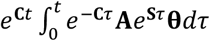 may contain a term linear in *t*. In which case we evaluate

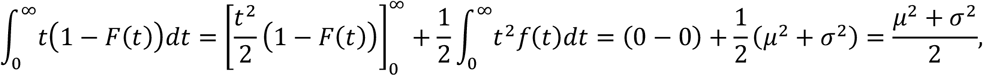

where *μ* and *σ* are the mean and standard deviation of the distribution for which *F* is the cumulative distribution function.

### Examples

#### No time-dependent removal hazard

For the model with removal hazard *h*(*t*) = 0, and thus *F*(*t*) = 0, it is convenient to return to the following expression for *R*_0_ within the derivation above:

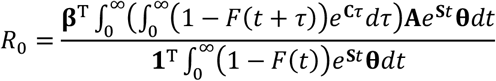

Plugging in *F*(*t*) = 0, we have

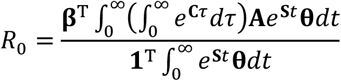

The following are true for invertible matrices **C** and **S**:

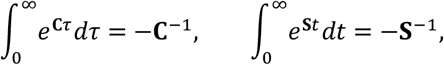

Hence

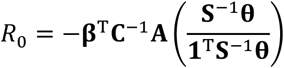

#### Model 1: Simple susceptible–colonized model

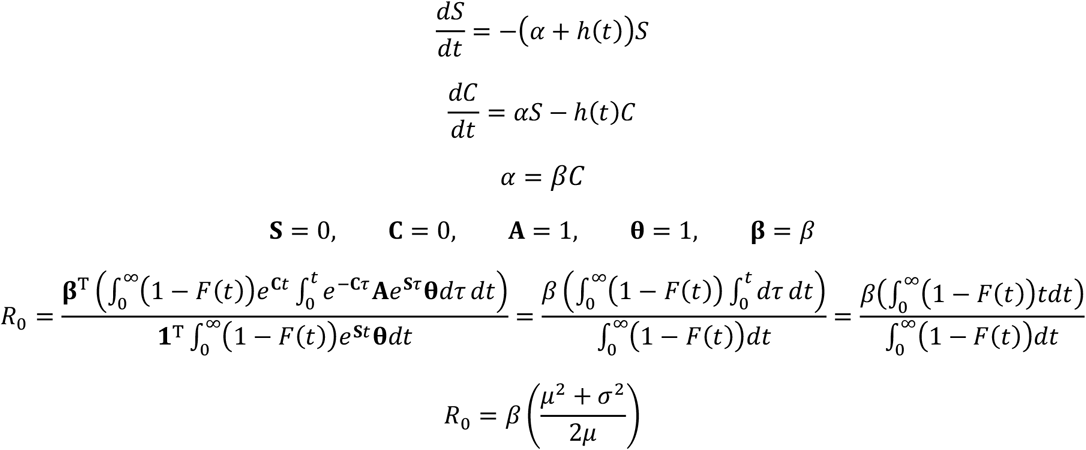

#### Model 2: Clearance of colonization

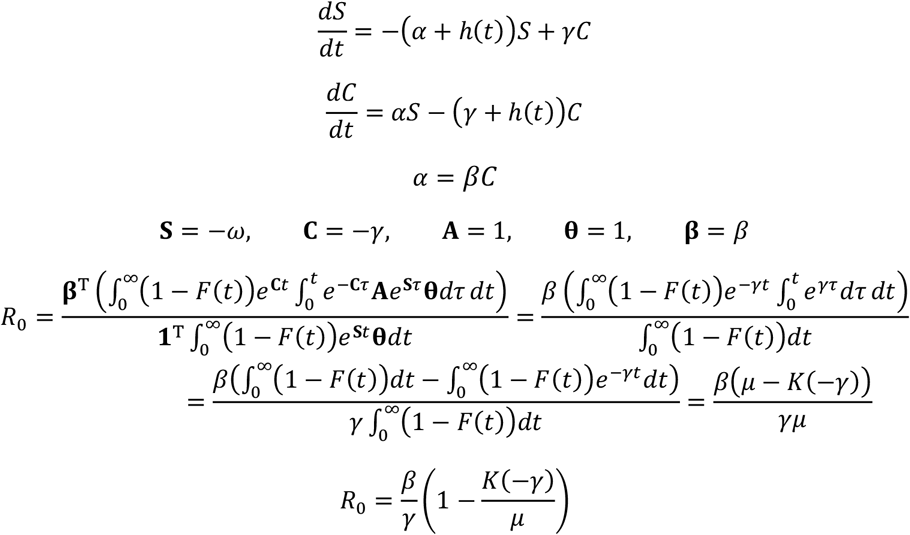

With length of stay exponentially distributed with rate *r*:

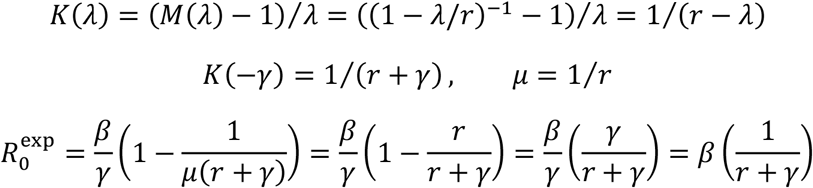

With length of stay gamma distributed with rate parameter r and shape parameter k

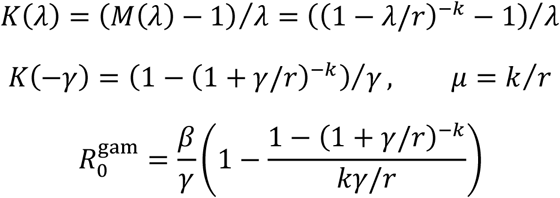

#### Model 3: Clinical detection with contact precautions

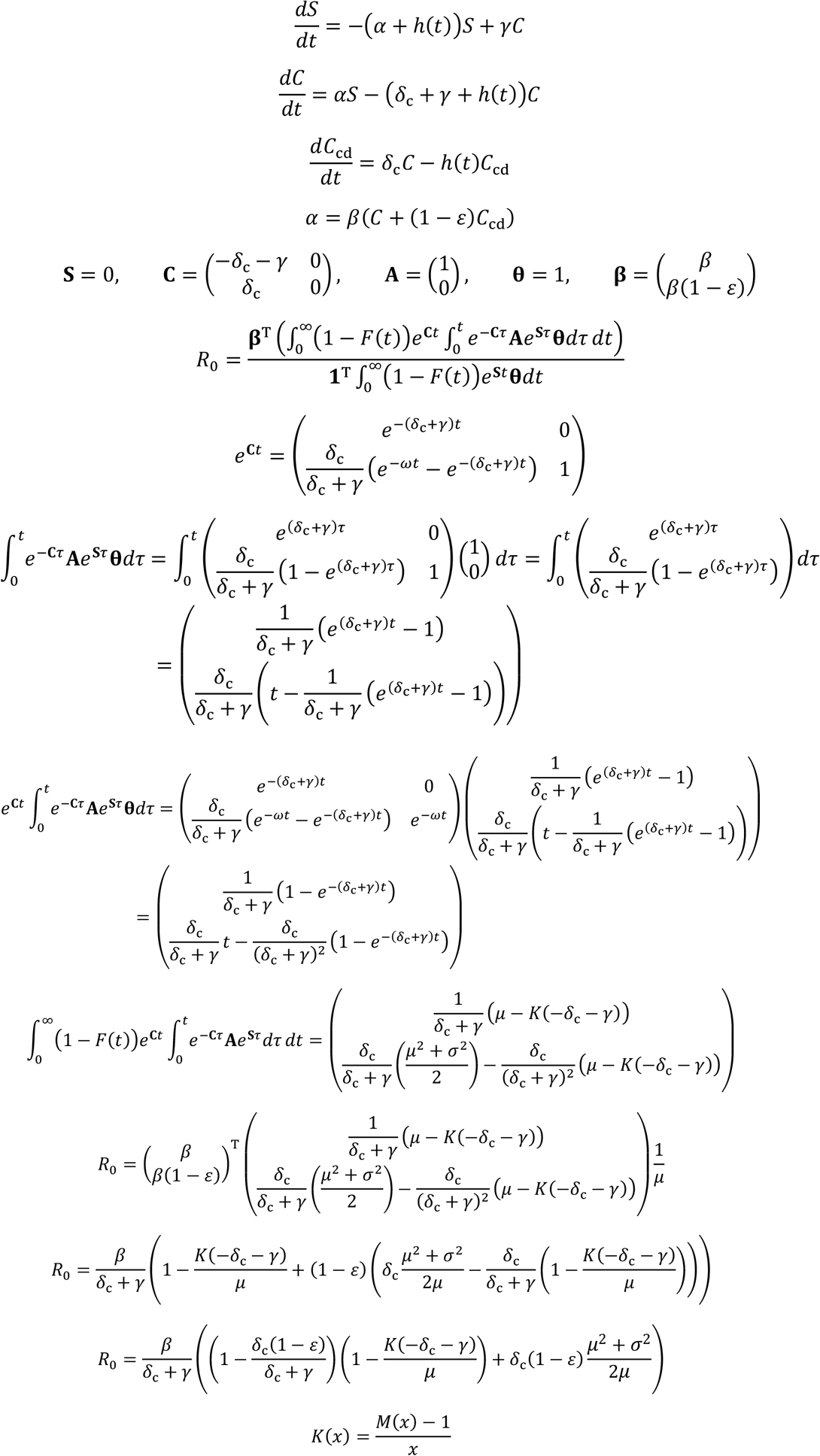

When applying this model to LTACH data, we assume a mixed exponential-gamma distribution for the length of stay, with a portion *p*_*x*_ of patients following an exponential distribution with mean *μ*_*x*_ and the rest following a gamma distribution with mean *μ*_*g*_ and shape parameter *k*. For this distribution:

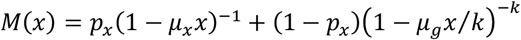

#### Model 4: Active surveillance and decolonization

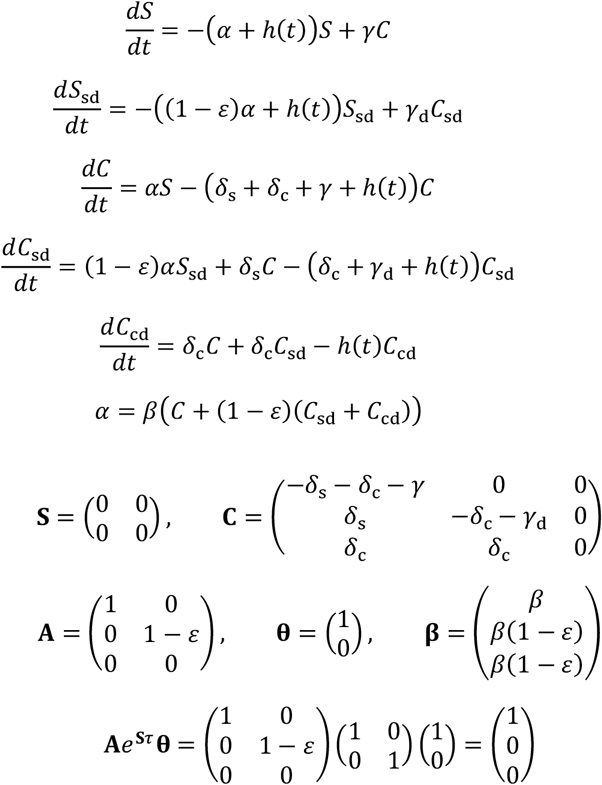

When *δ*_*s*_ **−** (*γ*_d_ **−** *γ*) ≠ 0:

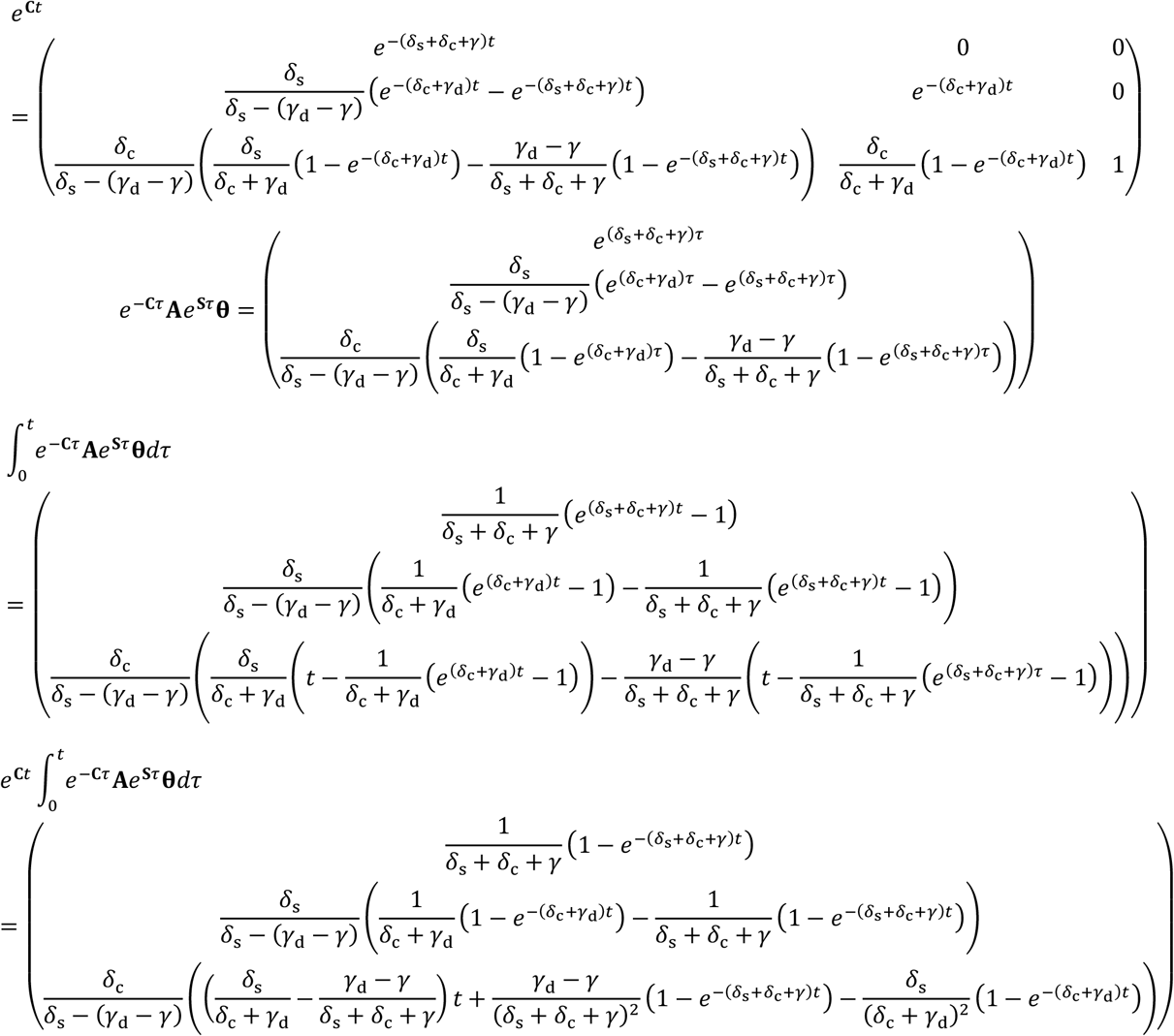

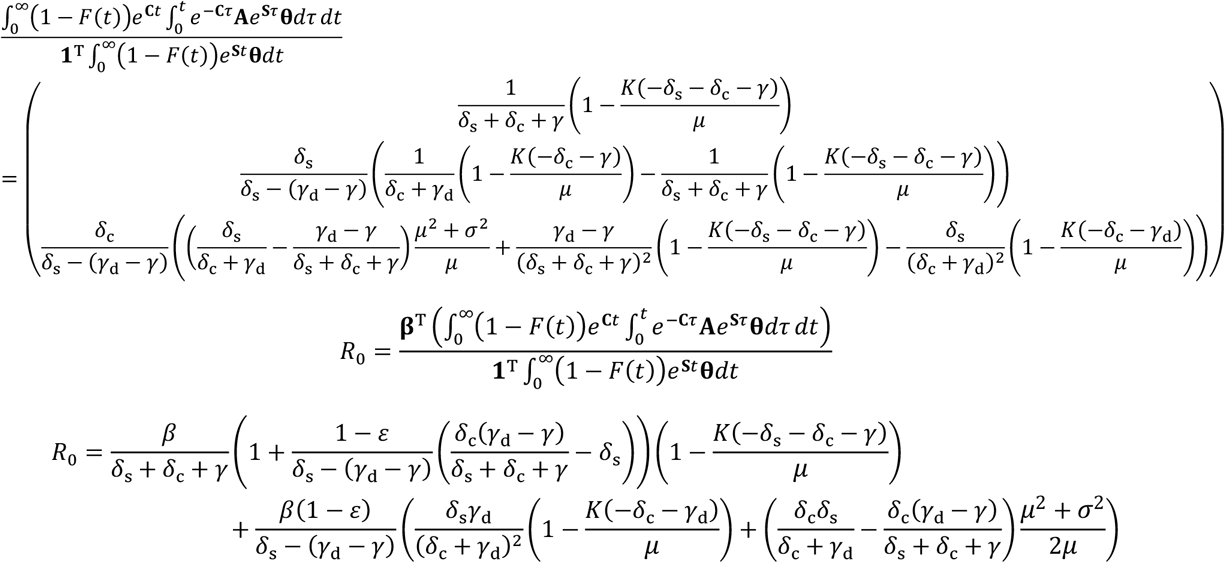

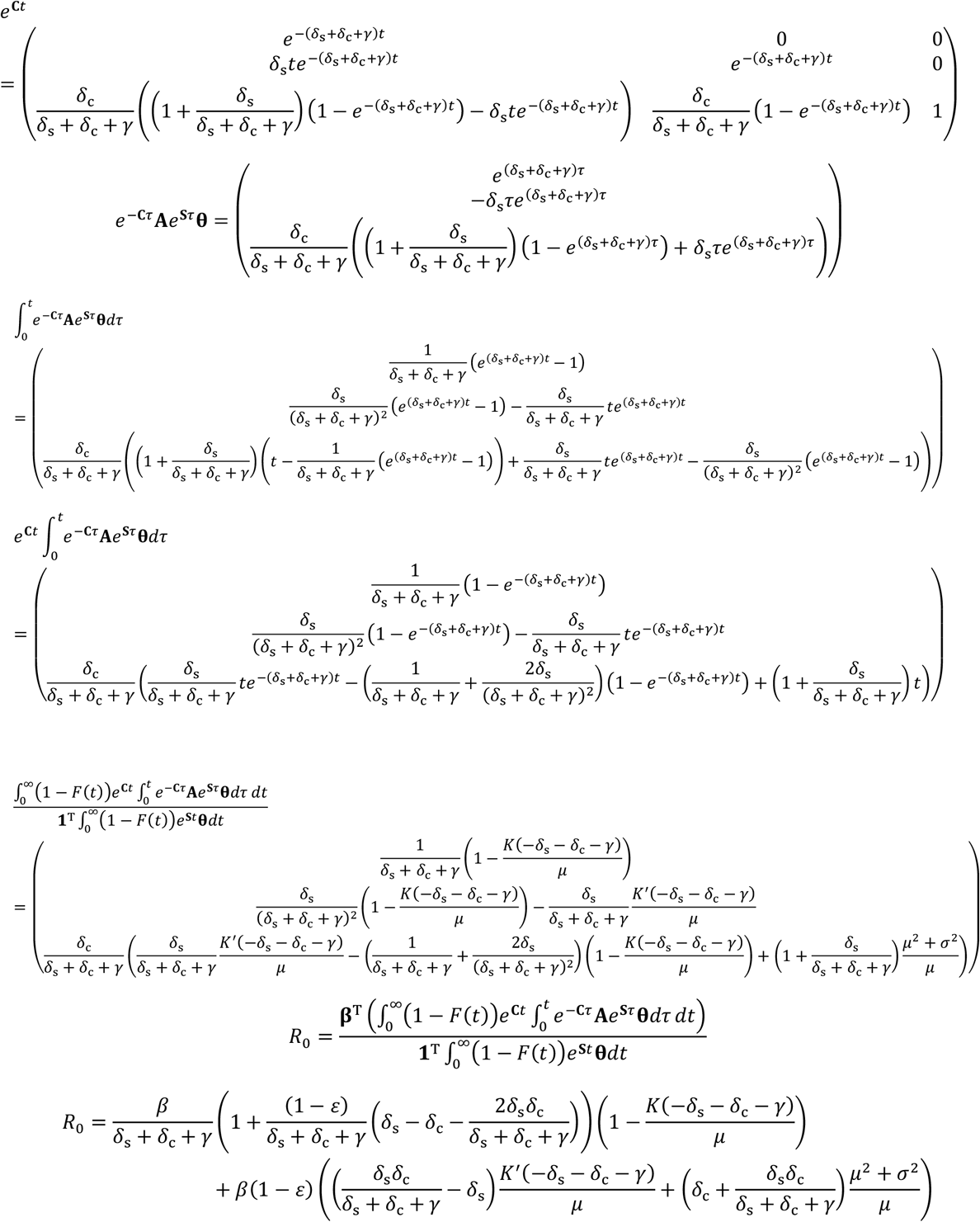

### Numerical calculation of R_0_

As demonstrated in the examples above, when the matrix exponentials e^**S***t*^ and e^**C***t*^ can be expressed symbolically for a particular model, the *R*_0_ formula we derived can be used directly to find a formula for *R*_0_ in terms of the model parameters. We can also calculate *R*_0_ numerically using the following procedure.

First, we create the following matrix **M**:

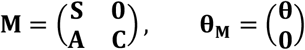

With **S**and **C** being *n × n* and *m × m* matrices, respectively, the **0** in the upper-right corner of **M** represents an *n* × *m* block of zeros. The **0** in the column vector *θ* **M** represents a column vector of *m* zeros.

The system

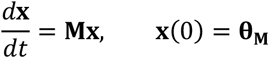

can be decoupled into two linked systems:

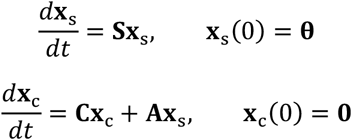

Solved sequentially we get:

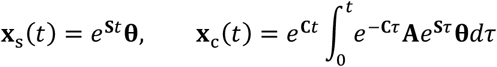

Returning to our *R*_0_ formula:

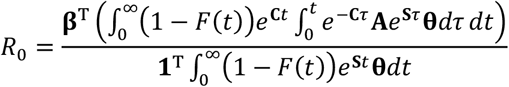

We can substitute in **x**_*s*_ and **x**_c_ to arrive at an alternate, equivalent expression:

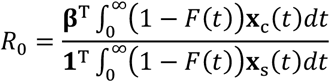

We can numerically calculate the numerator and denominator of that expression using the following procedure:

Numerically solve for the eigenvalues and eigenvectors of the matrix **M**. Place the eigenvalues in the diagonal matrix Λ and the eigenvectors in the corresponding columns of the matrix **V**, then calculate:

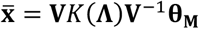

Then the integral 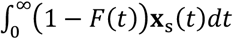 consists of elements 1 through *n* of 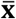, and 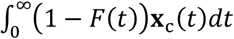 consists of elements *n* through *n* + *m* of 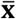.

The product **V**^**−**1^*θ*_**M**_ is calculated by solving the system **Vx** = *θ*_**M**_ for **x**, which does not require calculating **V**^**−**1^.

If the matrix **M** has repeated eigenvalues, the above procedure fails when there is an incomplete set of linearly independent eigenvectors with which to construct a nonsingular matrix **V**. In this case, our code attempts to calculate generalized eigenvectors to replace the duplicated eigenvectors in the matrix **V**. If **v**is an eigenvector of **M** associated with a repeated, degenerate eigenvalue *λ*, then a generalized eigenvector **g** solves the equation (**M −** *λ***I**)**g**=**v**.

To calculate a solution for **g** numerically, we first compute the Moore-Penrose inverse, also known as the pseudoinverse, (**M −** *λ* **I**)^+^, of the matrix (**M −** *λ* **I**). The Moore-Penrose inverse **B**^+^ of a matrix **B** satisfies **BB**^+^**B** = **B**. It can be shown that **g** = **B**^+^**v** is a solution to **Bg** = **v**. If we start with the equation to be solved and multiply both sides on the left by **BB**^+^, we get **BB**^+^**Bg** = **BB**^+^ **v**. Using the definitional property of **B**^+^, the left-hand side is equivalent to **Bg**, so **Bg** = **B**(**B**^+^**v**), which shows that **g** = **B**^+^ **v** solves the equation.

We use a numerical method for the singular value decomposition of the matrix (**M −** *λ* **I**) to calculate (**M −** *λ* **I**)^+^and then calculate a generalized eigenvector **g** = (**M −** *λ* **I**)^+^ **v**. Then, we construct a Jordan matrix **J** to replace Λ and a matrix **P** containing eigenvectors and generalized eigenvectors such that **M** = **PJP**^**−1**^. Then, the solution 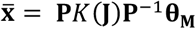, and the component *K*(**J**) includes elements above the diagonal involving derivatives of the function *K* evaluated at the repeated eigenvalue(s).

